# Clinical antiviral efficacy of remdesivir and casirivimab/imdevimab against the SARS-CoV-2 Delta and Omicron variants

**DOI:** 10.1101/2022.10.17.22281161

**Authors:** Podjanee Jittamala, William HK Schilling, James A Watson, Viravarn Luvira, Tanaya Siripoon, Thundon Ngamprasertchai, Pedro J Almeida, Maneerat Ekkapongpisit, Cintia Cruz, James J Callery, Simon Boyd, Orawan Anunsittichai, Maliwan Hongsuwan, Yutatirat Singhaboot, Watcharee Pagornrat, Runch Tuntipaiboontana, Varaporn Kruabkontho, Thatsanun Ngernseng, Jaruwan Tubprasert, Mohammad Yazid Abdad, Srisuda Keayarsa, Wanassanan Madmanee, Renato S Aguiar, Franciele M Santos, Elizabeth M Batty, Pongtorn Hanboonkunupakarn, Borimas Hanboonkunupakarn, Sakol Sookprome, Kittiyod Poovorawan, Mallika Imwong, Walter RJ Taylor, Vasin Chotivanich, Chunlanee Sangketchon, Wiroj Ruksakul, Kesinee Chotivanich, Sasithon Pukrittayakamee, Arjen M Dondorp, Nicholas PJ Day, Mauro M Teixeira, Watcharapong Piyaphanee, Weerapong Phumratanaprapin, Nicholas J White, the PLATCOV Collaborative Group

## Abstract

**Background:** Uncertainty over the therapeutic benefit provided by parenteral remdesivir in COVID-19 has resulted in varying treatment guidelines. Early in the pandemic the monoclonal antibody cocktail, casirivimab/imdevimab, proved highly effective in clinical trials but because of weak or absent *in vitro* activity against the SARS-CoV-2 Omicron BA.1 subvariant, it is no longer recommended.

**Methods:** In a multicenter open label, randomized, controlled adaptive platform trial, low-risk adult patients with early symptomatic COVID-19 were randomized to one of eight treatment arms including intravenous remdesivir (200mg followed by 100mg daily for five days), casirivimab/imdevimab (600mg/600mg), and no study drug. The primary outcome was the viral clearance rate in the modified intention-to-treat population derived from daily log_10_ viral densities (days 0-7) in standardized duplicate oropharyngeal swab eluates. This ongoing adaptive trial is registered at ClinicalTrials.gov (NCT05041907).

**Results:** Acceleration in mean estimated SARS-CoV-2 viral clearance, compared with the contemporaneous no study drug arm (n=64), was 42% (95%CI 18 to 73%) for remdesivir (n=67). Acceleration with casirivimab/imdevimab was 58% (95%CI: 10 to 120) in Delta (n=13), and 20% (95%CI: 3 to 43) in Omicron variant (n=61) infections compared with contemporaneous no study drug arm (n=84). In a *post hoc* subgroup analysis viral clearance was accelerated by 8% in BA.1 (95%CI: −21 to 59) and 23% (95%CI: 3 to 49) in BA.2 and BA.5 Omicron subvariants.

**Conclusions:** Parenteral remdesivir accelerates viral clearance in early symptomatic COVID-19. Despite substantially reduced *in vitro* activities, casirivimab/imdevimab retains *in vivo* antiviral activity against COVID-19 infections caused by currently prevalent Omicron subvariants.

**Brief summary:** In early symptomatic COVID-19 remdesivir accelerated viral clearance by 42% while the monoclonal antibody cocktail casirivimab/imdevimab accelerated clearance by approximately 60% in SARS-CoV-2 Delta variant infections, and by approximately 25% in infections with Omicron subvariants BA.2 and BA.5.

## Introduction

Remdesivir and the monoclonal antibody (mAb) cocktail, casirivimab/imdevimab, have both been used extensively as parenteral treatments for COVID-19, but therapeutic recommendations have varied widely. Early in the COVID-19 pandemic casirivimab/imdevimab proved highly effective in prevention and early treatment of COVID-19^1-5^, and showed life-saving efficacy in antibody negative hospitalized patients^6^. In contrast the initial evidence for parenteral remdesivir was unclear, and the WHO recommended against its use^7-15^, although this has recently changed^16,17^. Remdesivir is now widely recommended, whereas many regulatory authorities no longer recommend casirivimab/imdevimab. The various clinical trials on remdesivir and casirivimab/imdevimab were conducted largely in unvaccinated patients infected with the early SARS-CoV-2 viral variants which were more likely to result in hospitalization and severe outcomes than those prevalent today. These studies did not demonstrate an *in vivo* antiviral effect of remdesivir. In contrast the early casirivimab/imdevimab studies did show substantial acceleration in viral clearance, although they did not characterize the dose-response relationship *in vivo*. Governments around the world have spent approximately $17 billion on SARS-CoV-2 mAbs. This is greater than the nominal GDP of many countries. This year the USFDA^18^, the UK MHRA^19^, and now the WHO^20^, have recommended against use of casirivimab/imdevimab. These decisions were based upon substantially reduced activity in virus neutralization assays *in vitro* against the Omicron BA.1 subvariant^21-23^. No clinical assessments of activity had been reported, nor had *in vitro* SARS-CoV-2 antiviral activities been calibrated by *in vivo* data.

Meanwhile large randomized controlled trials (RCTs) have shown clinical efficacy in early COVID-19, and corresponding accelerated nasopharyngeal viral clearance, for both molnupiravir and nirmatrelvir (combined with ritonavir)^24-26^. None of these drugs have been compared directly. We present a randomized platform trial assessment of the *in vivo* antiviral activities of remdesivir and casirivimab/imdevimab in adults with acute symptomatic COVID-19 caused by the Delta and Omicron SARS-CoV-2 variants.

## Methods

PLATCOV is an ongoing phase 2 open label, randomized, controlled adaptive platform trial (ClinicalTrials.gov: NCT05041907) designed to provide a standardized quantitative comparative method for *in vivo* assessment of potential antiviral treatments in low-risk adults with early symptomatic COVID-19. The primary outcome measure is the viral clearance rate derived from the slope of the log_10_ oropharyngeal viral clearance curve over the first 7 days following randomization^27,28^. The treatment effect is defined as the multiplicative change in viral clearance rate relative to the no study drug arm. The trial was conducted in the Faculty of Tropical Medicine, Mahidol University, Bangkok; Bangplee hospital, Samut Prakarn; and Vajira hospital, Navamindradhiraj University, Bangkok, all in Thailand. The remdesivir arm was also conducted in testing centers in Belo Horizonte, Minas Gerais, Brazil (see Supplementary materials). All patients provided fully informed written consent. The PLATCOV trial was coordinated and monitored by the Mahidol Oxford Tropical Medicine Research Unit (MORU) in Bangkok, was overseen by a trial steering committee (TSC) and its results were reviewed regularly by a data and safety monitoring board (DSMB). The funders had no role in the design, conduct, analysis or interpretation of the trial.

### Participants and procedures

Previously healthy adults aged between 18 and 50 years were eligible for the trial if they had early symptomatic COVID-19 (i.e. reported symptoms for ≤4 days), oxygen saturation ≥96%, were unimpeded in activities of daily living, and gave fully informed consent. SARS-CoV-2 positivity was defined either as a nasal lateral flow antigen test which became positive within two minutes (STANDARD™ Q COVID-19 Ag Test, SD Biosensor, Suwon-si, Korea) or a positive PCR test within the previous 24hrs with a cycle threshold value (Ct) <25 (all viral gene targets), both suggesting high viral loads. Exclusion criteria included taking any potential antivirals or pre-existing concomitant medications, chronic illness or significant comorbidity, hematological or biochemical abnormalities, pregnancy (a urinary pregnancy test was performed in females), breastfeeding, or contraindication or known hypersensitivity to any of the study drugs.

Enrolled patients were either admitted to the study ward (in Thailand) or followed as outpatients at home (in Brazil). After randomization and baseline procedures (see Supplementary materials) oropharyngeal swabs (two swabs from each tonsil) were taken as follows. A flocked swab (Thermo Fisher MicroTest™ and later COPAN FLOQSwabs^®^) was rotated against the tonsil through 360° four times and placed in Thermo Fisher M4RT™ viral transport medium (3mL). Swabs were transferred at 4-8°C, aliquoted and then frozen at −80°C within 48hrs. Separate swabs from each tonsil were taken once daily from day 0 to day 7, and on day 14. Vital signs were recorded three times daily and symptoms and any adverse effects were recorded daily.

The TaqCheck™ SARS-CoV-2 Fast PCR Assay (Applied Biosystems^®^, Thermo Fisher Scientific, Waltham, Massachusetts) quantitated viral loads (RNA copies per mL). This multiplexed real-time PCR method detects the SARS-CoV-2 N and S genes, and human RNase P in a single reaction. RNase P helped correct for variation in sample human cell content. Viral loads were quantified against ATCC^®^ heat-inactivated SARS-CoV-2 (VR-1986HK™ strain 2019-nCoV/USA-WA1/2020) standards. Viral variants were identified using Whole Genome Sequencing (Supplementary materials). Adverse events were graded according to the Common Terminology Criteria for Adverse Events v.5.0 (CTCAE). Summaries were generated if the adverse event was ≥grade 3 and was new or had increased in intensity. Serious adverse events were recorded separately and reported to the DSMB.

### Outcome measures and statistical analysis

The primary outcome measure was the rate of viral clearance, expressed as a slope coefficient (28), and estimated under a Bayesian hierarchical linear model fitted to the daily log_10_ viral load measurements between days 0 and 7 (18 measurements per patient). The viral clearance rate (i.e. slope coefficient from the model fit) can be expressed as a clearance half-life (t_1/2_ = log_10_ 0.5/slope). The treatment effect was defined as the multiplicative change (%) in the viral clearance rate relative to the no study drug arm (i.e. how much the test treatment accelerates viral clearance)^27^. A 50% increase in clearance rate thus equals a 33% reduction in clearance half-life. All cause hospitalization for clinical deterioration (until day 28) was a secondary endpoint. For each studied intervention the sample size is adaptive based on prespecified futility and success stopping rules.

All analyses were done in a modified intention-to-treat (mITT) population, comprising patients who had ≥3 days follow-up data. A series of linear and non-linear Bayesian hierarchical models were fitted to the viral quantitative PCR (qPCR) data (Supplementary materials). Model fits were compared using approximate leave-one-out comparison as implemented in the package *loo*. All data analysis was done in R version 4.0.2. Model fitting was done in *stan* via the *rstan* interface. Model fits were compared using approximate leave-one-out comparison as implemented in the package *loo*. All code and data are openly accessible via GitHub: https://github.com/jwatowatson/PLATCOV-Remdesivir-Regeneron.

## Results

The trial began recruitment on 30 September 2021. On 10 June 2022, remdesivir enrolment was stopped as the prespecified success margin had been reached. Of the 439 patients screened by that time, 337 were randomized to either remdesivir (67 patients), no study drug (69 patients), or to other interventions (201 patients: casirivimab/imdevimab, ivermectin, favipiravir, nitazoxanide, fluoxetine, molnupiravir, or nirmatrelvir/ritonavir). On 24 August 2022, casirivimab/imdevimab was unblinded because the prespecified success margin had been reached. This report includes patients randomized to casirivimab/imdevimab or no study drug arm up until unblinding. Of the 528 patients screened, 74 were randomized to casirivimab/imdevimab; 89 to no study drug; and 269 to other interventions. Five patients were excluded from both the remdesivir and casirivimab/imdevimab analyses (Figure 1), resulting in mITT populations of n=131 (67 remdesivir and 64 no study drug) and n=158 (74 casirivimab/imdevimab and 84 no study drug).

**Figure 1:**
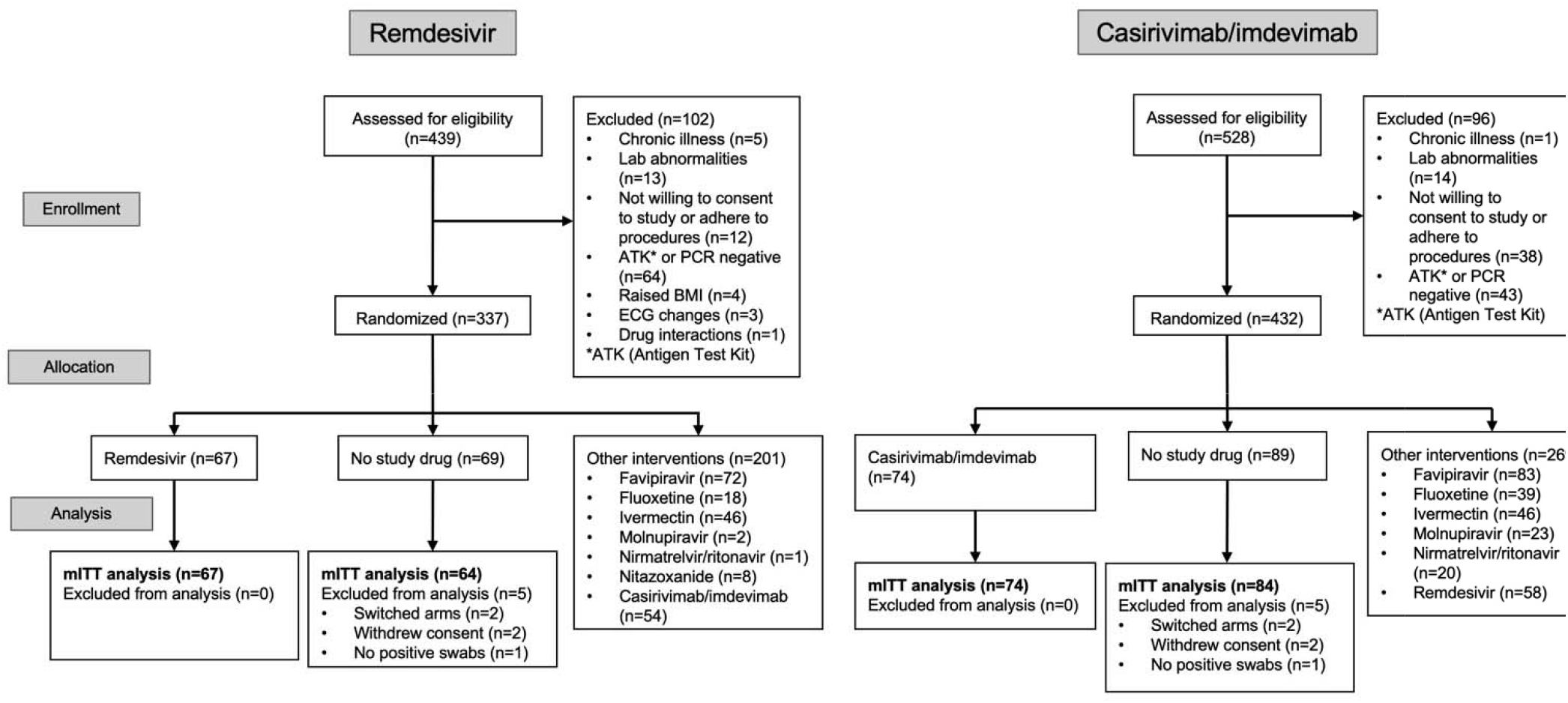
CONSORT diagram of the PLATCOV phase 2 open label, randomized, controlled adaptive platform trial for remdesivir (Thailand and Brazil. Enrolment end: 10 June 2022) and casirivimab/imdevimab (Thailand only. Enrolment end: 24 August 2022)

### Virological responses

All analytical models of oropharyngeal virus clearance were in excellent agreement, although the non-linear model gave smaller effect size estimates (see Supplementary figures). In general, the non-linear model (which allows some patients to have viral load increases after randomization) fitted the data better. All effect sizes reported in the main text are inferred under the main model: a linear model with intercept and slope adjusted for site and virus variant.

### Remdesivir

The 131 patients in the remdesivir analysis mITT population had a median of 18 viral load measurements (range 16-18) each between days 0 and 7, of which 14.6% (343/2,356) were below the lower limit of detection (LOD). The baseline geometric mean (GM) oropharyngeal viral load was 3.5×10^5^ RNA copies/mL (IQR 5.9×10^4^ to 2.5×10^6^). Relative to the no study drug arm, clearance of oropharyngeal virus in patients randomized to remdesivir was 42% faster (95%CI 18% to 73%; probability of >12.5% acceleration: 0.99) (Figures 2 and 3). The median estimated viral clearance half-lives under the linear model were 12.8 (range: 4.8-50.0) hours in the remdesivir arm and 18.0 (range: 3.6-46.7) hours in the no study drug arm, corresponding with a shortening of the median virus clearance half-life by approximately one third.

**Figure 2:**
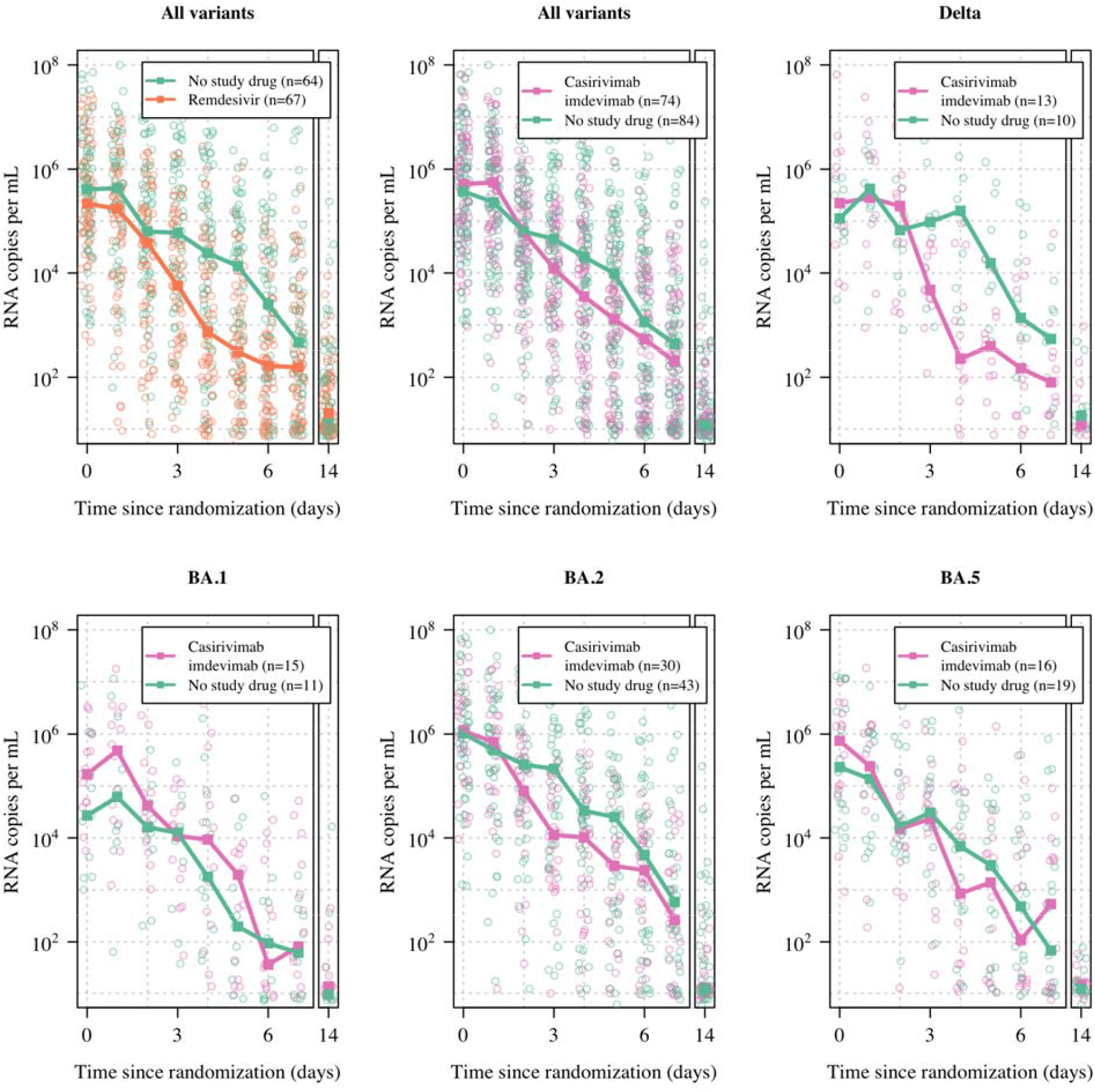
Median SARS-CoV-2 oropharyngeal virus clearance profiles following remdesivir (orange), casirivimab/imdevimab (purple) and no study drug (green) arms. For casirivimab/imdevimab we show clearance profiles by major variants. BA.4 is not shown as there was only one patient with this variant (in the no study drug arm).

**Figure 3.**
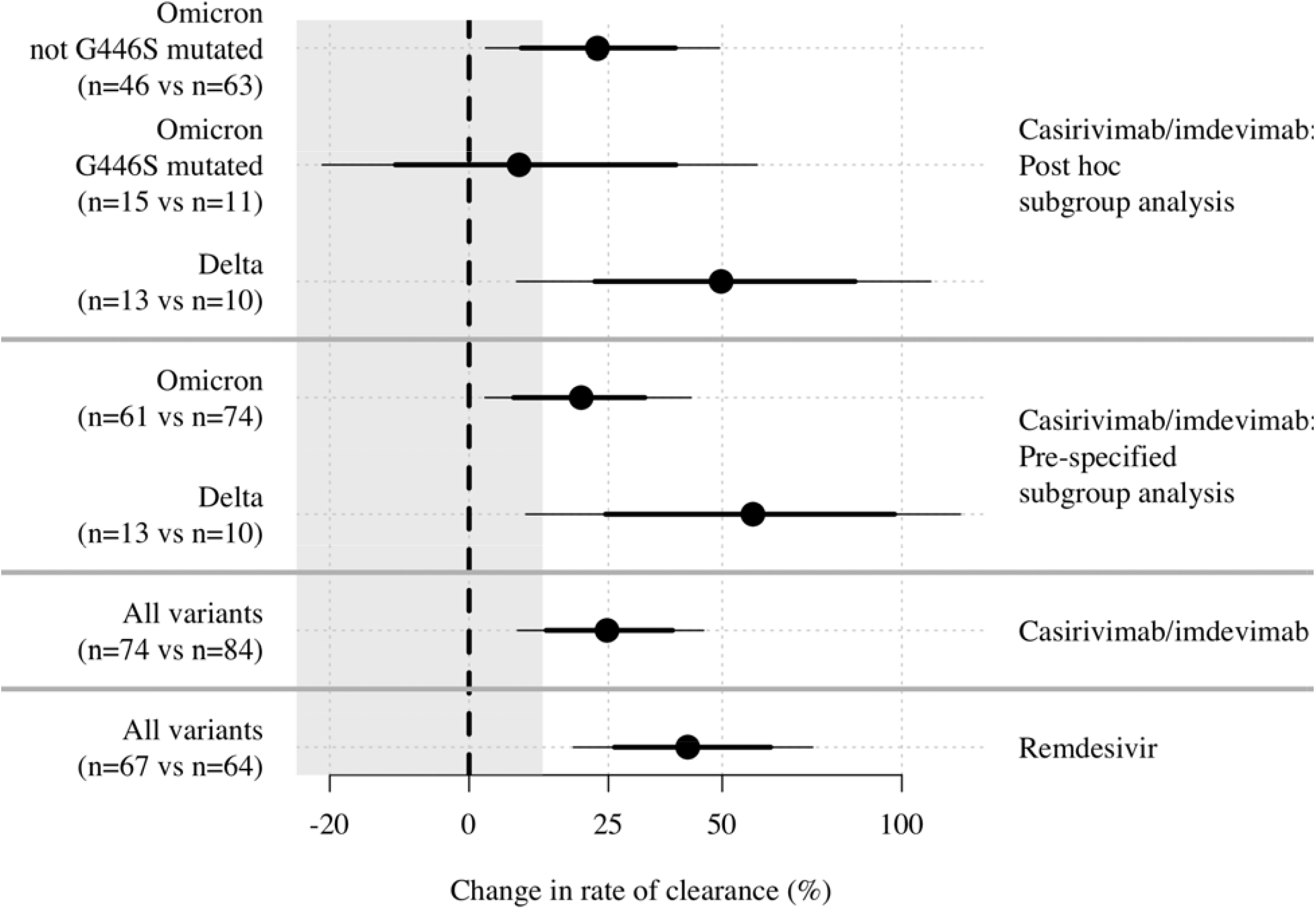
Estimated treatment effect of intravenous remdesivir and the casirivimab/imdevimab monoclonal antibody cocktail in acute early COVID-19. Mean posterior estimates of the change in the rate of viral clearance (black circles) compared to the contemporaneous no study drug arm. The 80% and 95% credible intervals are shown by the thick and thin lines, respectively. The thick grey horizontal lines separate the different analyses.

### Casivirimab/imdevimab

Of the 18 individual viral load measurements, 11.7% (331/2,842) from 158 patients in the casirvimab/imdevimab analysis mITT population were below the LOD. The baseline geometric mean oropharyngeal viral load was 3.8×10^5^ RNA copies/mL (IQR 2.0×10^3^ to 7.2×10^7^), (Figure 2). Overall, viral loads declined 25% faster (95%CI 8 to 46) in casirivimab/imdevimab recipients compared to the no study drug arms (Figure 3). However, there was substantial heterogeneity in the treatment effect for the different virus variants. In the primary pre-specified subgroup analysis, under an interaction model, viral clearance rates in Delta variant infections were increased by 58% (95% CI: 10 to 120%) relative to no study drug. In the Omicron variants overall, this effect was reduced by nearly three-fold: 20% (95% CI: 3 to 43%); posterior probability of less effect in Omicron compared with Delta: 0.91 (Figure 3).

### Genotype associations

In laboratory studies casirivimab has no significant *in vitro* activity against Omicron variants^21-23^. For imdevimab mutations (N440K and G446S) in the ACE2 receptor binding site cause reduced *in vitro* neutralizing activity^21-23,29-31^. N440K occurs in all Omicron subvariants and 2-3log decrease in *in vitro* neutralization whereas G446S, which causes a substantially larger reduction in *in vitro* activity, occurs only in BA.1, BA.2.75 and BA.3 lineages. In a secondary *post hoc* subgroup analysis, we estimated treatment effects across Delta, Omicron G446S mutated (all BA.1), and Omicron G446 wild-type (BA.2, BA.4 and BA.5) variants. Under an interaction model we estimated a 50% increase in viral clearance rate for Delta (95% CI: 8 to 109); an 8% increase for BA.1 (95%CI: −21 to 59%); and a 23% increase in viral clearance rate for Omicron variants which were not G446S mutated (95%CI: 3 to 49%) (Figure 3).

### All data combined

Analyzing data in the two combined mITT populations under a single model (n=232), infections with the Omicron subvariants BA.2 and BA.5 cleared slightly faster than the Delta variant. Patients with BA.2 had higher baseline viral loads (1.47-fold higher, 95%CI 1.04 to 2.03), (Supplementary Results). The interval since symptom onset was a strong predictor of the baseline viral load (0.65 fold lower per additional day, 95%CI 0.54-0.76). Under the full model, casirivimab/imdevimab in the Delta variant had the greatest effect on acceleration of viral clearance (probability 0.90), followed by remdesivir (probability 0.81) and then casirivimab/imdevimab in the Omicron variants (probability 0.89). There was one hospitalization in the no study drug arm (see below).

### Adverse effects

The oropharyngeal swabbing and all treatments were well tolerated. There were three serious adverse events (SAE) in the no study drug arm, and one in the remdesivir arm. Two patients in the no study drug arm and one in the remdesivir arm had asymptomatic raised creatinine phosphokinase (CPK) level (>10 times ULN) attributed to COVID-19-related skeletal muscle damage. This improved with fluids and supportive management and was considered unrelated to treatment. One patient (no study drug arm) was readmitted one day after discharge because of chest pain and lethargy. All clinical and laboratory investigations were normal and the patient was discharged the following day. Three patients reported a rash with casirivimab/imdevimab, which were not SAEs and resolved spontaneously. There were no treatment related serious adverse events.

## Discussion

This comparative *in vivo* pharmacodynamic assessment of parenteral therapeutics in early COVID-19 infections shows that both remdesivir and casirivimab/imdevimab accelerate viral clearance but, for the monoclonal antibody, the effect depended on the SARS-CoV-2 variant. Continued uncertainty over the value of different COVID-19 treatments has resulted in substantial variation in therapeutic guidelines and clinical practice. Until 22 April 2022, because of unclear benefits in hospitalized patients, the World Health Organization ‘living guideline’ gave a conditional recommendation against remdesivir^15^. Many countries have not licensed remdesivir for use. However, it is increasingly clear that antivirals are more effective in early COVID-19, when viral burdens are highest, than in late-stage disease (i.e. hospitalized patients requiring oxygen or ventilation) where viral burdens have declined and anti-inflammatory interventions are more effective^32,33^. The average 42% increase in viral clearance rate observed with remdesivir is similar in magnitude to that observed in trials with molnupiravir, although the assessment methodologies were different^24,26^. The substantial intra-individual variability in nasopharyngeal (or oropharyngeal) viral load estimates results in a low signal to noise ratio^27^ which explains why studies which sampled infrequently, such as PINETREE^16^, failed to associate increased viral clearance with therapeutic efficacy.

Based upon *in vitro* testing of the Omicron BA.1 variant, some regulatory authorities have withdrawn use authorization for casirivimab/imdevimab^18,19^. This pharmacometric study indicates strong activity of casirivimab/imdevimab in SARS-CoV-2 Delta variant infections similar to that reported against early variants in the preregistration studies^2,5^ but, contrary to general opinion, the antibody cocktail retains measurable *in vivo* activity against currently prevalent Omicron subvariants – presumably because of the partial activity of imdevimab. All Omicron subvariants contain mutations which abolish casirivimab activity but many are still susceptible to imdevimab. Our *post hoc* subgroup analysis, informed by recent detailed in-vitro assessments^34^, was compatible with little or no *in vivo* activity against BA.1 (as expected from the *in vitro* studies), but showed efficacy against the BA.2 and BA.5 variants. Nearly all Omicron subvariants retain the N440K mutation in the spike protein ACE2 receptor binding motif associated with reduced imdevimab activity *in vitro*. The BA.1 subvariant also usually has the G446S mutation which largely abolishes imdevimab *in vitro* activity (IC_50_ ≥10,000ng/mL)^21-23^. But in most BA.2, BA.4 and BA.5 sub-lineages, the G446S mutation has reverted back to wild-type. The resulting moderate in-vitro activity in most BA.2, BA.4 and BA.5 sub-lineages was presumed to be clinically insignificant. But the doses, and thus *in vivo* exposures of therapeutic antibodies are very high in relation to *in vitro* potency^34^. Many *in vitro* studies evaluated activities only in the nanomolar range, and did not investigate whether maximal effects were achievable at micromolar concentrations^21-23^ which are readily achieved in-vivo^5^. The methodology used to assess *in vitro* neutralization, and the reported inhibitory concentrations also vary widely^29,34^. The approximate 23% acceleration in viral clearance with casirivimab/imdevimab in COVID-19 infections observed with prevalent Omicron subvariants is less than observed earlier in Delta variant infections, and there are other agents which are probably more active (if they can be accessed and afforded). But as further spike protein evolution could negate this activity^34^, the time-critical question now is whether this level of antiviral activity is clinically useful, and thus whether the use authorizations should be reinstated and the recommendations against use reversed?

The main limitation of our study is the small numbers in each of the major SARS-CoV-2 sub-lineages limiting the confidence of our estimates of the relative effects of casirivimab/imdevimab. In particular there were few BA.1 infections, so we could not make a precise treatment effect estimate. Another important limitation is that this is an open label study, which led to more withdrawals in the no study drug arm.

This simple pharmacometric methodology is readily performed anywhere with accurate qPCR viral quantitation. Duplicate daily oropharyngeal swabs are well tolerated (whereas daily nasopharyngeal swabbing is not). It provides a rapid assessment of relative antiviral efficacy and so can be used to characterize dose-response relationships in real time and thereby inform therapeutic practice. Regulatory authority and treatment guideline decisions should be based upon *in vivo*, as well as *in vitro* evidence^34^. The very large amounts of casirivimab/imdevimab which have been purchased, and will soon reach their expiry dates, could still provide therapeutic benefit for high-risk patients with COVID-19. Many countries currently have no access to alternative effective medicines.

## Supporting information

Master Protocol

Latest Statistical Analysis Plan

## Data Availability

All code and data are openly accessible via GitHub: https://github.com/jwatowatson/PLATCOV-Remdesivir-Regeneron.

## Acknowledgements

We thank all the patients with COVID-19 who volunteered to be part of the study. We thank the data safety and monitoring board (DSMB) (Tim Peto, Andre Siqueira, and Panisadee Avirutnan); the trial steering committee (TSC) (Nathalie Strub-Wourgaft, Martin Llewelyn, Deborah Waller, and Attavit Asavisanu); Sompob Saralamba and Tanaphum Wichaita for developing the RShiny randomization app; and Mavuto Mukaka for invaluable statistical support. We also thank all the staff of the Clinical Trials Unit (CTU) at MORU, Shivani Singh for research into use and purchasing of monoclonal antibodies, Praram 9 Hospital Bangkok for selling at a discounted price some of the doses of casirivimab/imdevimab, PCR Expert group (Janjira Thaipadungpanit, Audrey Dubot-Pérès and Clare Ling), Thermo Fisher for their excellent support with this project, and all the hospital staff at the Hospital of Tropical Diseases (HTD), Bangplee (BP) and Vajira (VJ) hospitals, as well as those involved in sample processing in MORU and the processing and analysis at the Faculty of Tropical Medicine, molecular genetics laboratory. We would thank the MORU Clinical Trials Support Group (CTSG) for data management and logistics, and the purchasing, administration and support staff at MORU, and those at the Brazil site who provided expert help in managing patients (Joseane Fratari, Josiane Vaz, Fátima Brant and Lísia Esper).

“Finding treatments for COVID-19: A phase 2 multi-centre adaptive platform trial to assess antiviral pharmacodynamics in early symptomatic COVID-19 (PLAT-COV)” is supported by the Wellcome Trust Grant ref: 223195/Z/21/Z through the COVID-19 Therapeutics Accelerator.

---

**Table 1.**
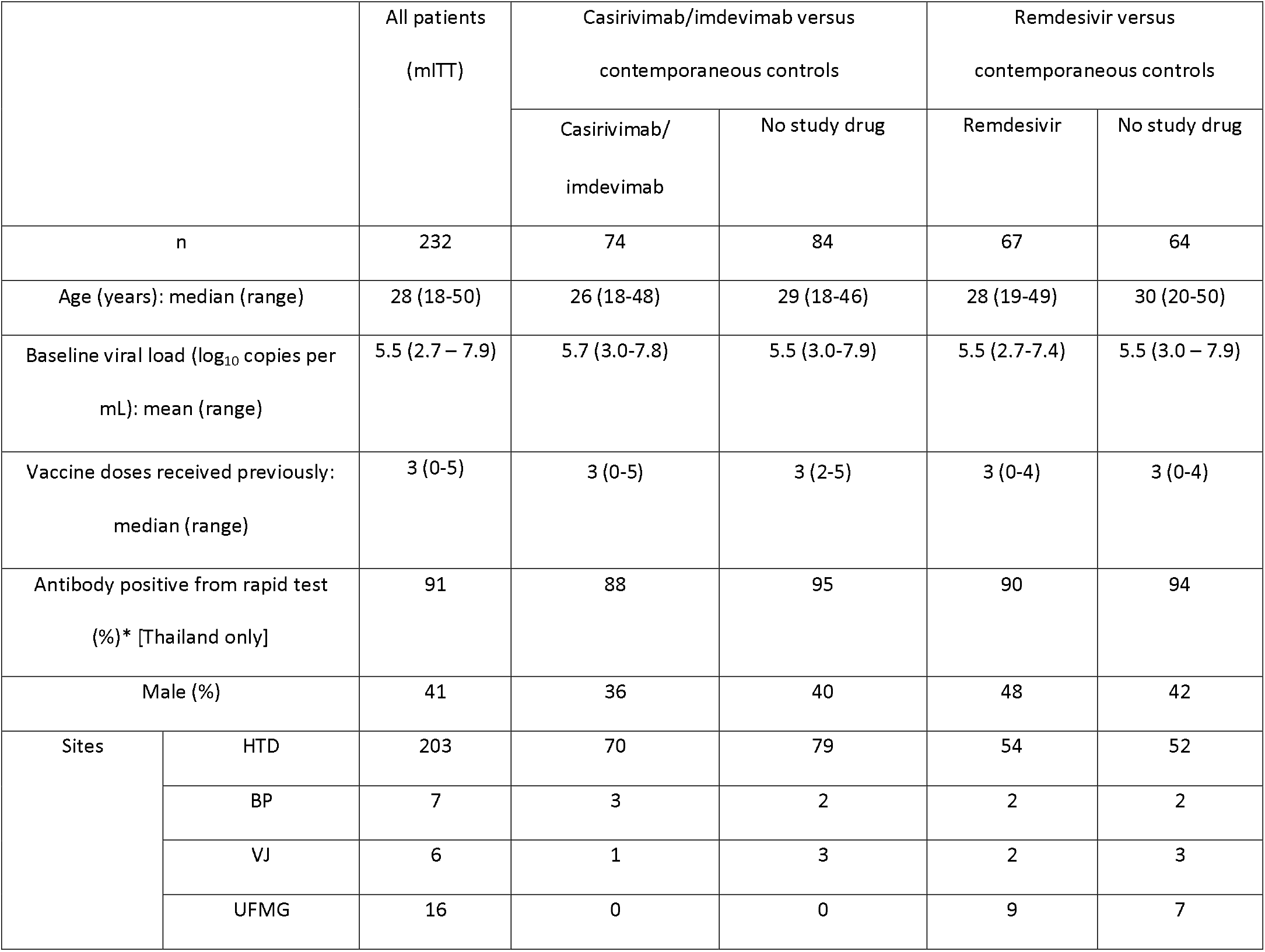
Summary of patient characteristics included in the two mITT populations (total n=232). HTD: Hospital for Tropical Diseases, Thailand; BP: Bangplee Hospital, Thailand; VJ: Vajira Hospital, Thailand; UFMG: Universidade Federal de Minas Gerais, Brazil. *Defined as IgM or IgG present on the rapid antibody test (BIOSYNEX COVID-19 BSS IgM/IgG, Illkirch-Graffenstaden, France) used as per manufacturer’s instructions

