## Supplementary materials for "Clinical antiviral efficacy of remdesivir and casirivimab/imdevimab against the SARS-CoV-2 Delta and Omicron variants"

### List of Sites and Investigators (PLATCOV Collaborative Group)

**Sites**

1. Hospital for Tropical Diseases (HTD), Faculty of Tropical Medicine, Mahidol University, 420/6 Rajvithi Road, Bangkok, 10400, Thailand
2. Vajira Hospital (VJ), Navamindradhiraj University, 681 Samsen road, Dusit, Bangkok, 10300, Thailand
3. Bangplee Hospital (BP), 88/1 Moo 8 Tambon Bang Phli Yai, Amphoe Bangplee, Samut Prakan 10540, Thailand
4. Universidade Federal de Minas Gerais, Av. Antônio Carlos, 6627 Belo Horizonte, Minas Gerais 31270 – 901, Brazil

**Investigators**

Co-principal investigators:

Nicholas J White^1,2^

William HK Schilling^1,2^

Faculty of Tropical Medicine:

Site and Country Principal investigator:

Weerapong Phumratanaprapin^4^

Accountable Investigator:

Viravarn Luvira^4^

Co-Investigators:

James J Callery^1,2^

Nicholas PJ Day^1,2^

Sasithon Pukrittayakamee^1,4^

Simon Boyd^1,2^

Cintia Cruz^1,2^

Arjen M Dondorp^1,2^

Walter RJ Taylor^1,2^

James A Watson^1,2^

Watcharapong Piyaphanee^4^

Kittiyod Poovorawan^1,4^

Thundon Ngamprasertchai^4^

Tanaya Siripoon^4^

Borimas Hanboonkunupakarn^1,4^

Kesinee Chotivanich^1,4^

Podjanee Jittamala^1,3^

Mallika Imwong^1,5^

Maneerat Ekkapongpisit^1^

Varaporn Kruabkontho^1^

Thatsanun Ngernseng^1^

Jaruwan Tubprasert^1^

Mohammad Yazid Abdad^1,2^

Srisuda Keayarsa^4^

Orawan Anunsittichai^1^

Maliwan Hongsuwan^1^

Yutatirat Singhaboot^4^

Wanassanan Madmanee^1^

Elizabeth M Batty^1,2^

Runch Tuntipaiboontana^1^

Watcharee Pagornrat^1^

Vajira Hospital:

Site Principal investigator:

Vasin Chotivanich^7^

Co-investigators:

Wiroj Ruksakul^7^

Chunlanee Sangketchon^8^

Bangplee Hospital:

Site Principal investigator:

Pongtorn Hanboonkunupakarn^6^

Co-investigator:

Sakol Sookprome^6^

Brazil Site (Universidade Federal de Minas Gerais):

Site and Country Principal investigator:

Mauro M Teixeira^9^

Co-Investigators:

Pedro J Almeida^9^

Renato S Aguiar^10^

Franciele M Santos^10^

**Affiliations:**

1. Mahidol Oxford Tropical Medicine Research Unit, Faculty of Tropical Medicine, Mahidol University, Bangkok, Thailand

2. Centre for Tropical Medicine and Global Health, Nuffield Department of Medicine, Oxford University, Oxford, UK

3. Department of Tropical Hygiene, Faculty of Tropical Medicine, Mahidol University, Bangkok, Thailand

4. Department of Clinical Tropical Medicine, Faculty of Tropical Medicine, Mahidol University, Bangkok, Thailand

5. Department of Molecular Tropical Medicine and Genetics, Faculty of Tropical Medicine, Mahidol University, Bangkok, Thailand

6. Bangplee Hospital, Ministry of Public Health, Thailand

7. Faculty of Medicine, Navamindradhiraj University, Bangkok,Thailand

8. Faculty of Science and Health Technology, Navamindradhiraj University, Bangkok,Thailand

9. Clinical Research Unit, Center for Advanced and Innovative Therapies, Universidade Federal de Minas Gerais, Brazil

10. Department of Genetics, Ecology and Evolution, Institute of Biological Sciences, Universidade Federal de Minas Gerais

### Ethics Approval

The trial was approved by local and national research ethics boards in Thailand (Faculty of Tropical Medicine Ethics Committee, Mahidol University, FTMEC Ref: TMEC 21-058) and the Central Research Ethics Committee (CREC, Bangkok, Thailand, CREC Ref: CREC048/64BP-MED34), in Brazil by the Research Ethics Committee of the Universidade Federal de Minas Gerais (COEP-UFMG, Minas Gerais, Brazil, COEP-UFMG) and National Research Ethics Commission- (CONEP, Brazil, COEP-UFMG and CONEP Ref: CAAE:51593421.1.0000.5149), and by the Oxford University Tropical Research Ethics Committee (OxTREC, Oxford, UK, OxTREC Ref: 24-21).

### Baseline procedures

Baseline investigations included a full clinical examination, rapid SARS-CoV-2 antibody test (BIOSYNEX COVID-19 BSS IgM/IgG®, Illkirch-Graffenstaden, France, done in Thailand only), blood sampling for hematology and biochemistry, an electrocardiogram and a chest radiograph (following local guidance in Thailand, but not a study requirement).

### Remdesivir and casirivimab/imdevimab additional information

Remdesivir- Covifor™ (Hetero Drugs Ltd., Hyderabad, India) in Thailand (n=58), and Veklury™ (Gilead Sciences, Inc. Foster City, California) in Brazil (n=9), was given by rate-controlled infusion (reconstituted and added to 250mls 0.9% saline and given over 60 minutes) in an initial adult dose of 200mg, and then 100mg infusion once daily over the next four days to complete a five day course.

Casirivimab/imdevimab (600mg/600mg; Roche, Switzerland) was given once by intravenous infusion following randomization.

### Adverse events (AE) for remdesivir

Supplementary table 1: Summary of adverse events (grade 3 and above) for remdesivir

|  | **All grades** | | **Grade 3-4** | |
| --- | --- | --- | --- | --- |
|  | **Remdesivir**  **(n=67)** | **No study drug (n=69)** | **Remdesivir**  **(n=67)** | **No study drug (n=69)** |
| Any adverse event (Grade ≥ 3) | 1 | 2 |  |  |
| Serious adverse event reported | 1 | 2 |  |  |
| Symptoms |  |  |  |  |
| Fever |  |  | 0 | 0 |
| Headache |  |  | 0 | 0 |
| Dizziness |  |  | 0 | 0 |
| Blurred vision |  |  | 0 | 0 |
| Fatigue |  |  | 0 | 0 |
| Cough |  |  | 0 | 0 |
| Difficulty breathing |  |  | 0 | 0 |
| Chest pain |  |  | 0 | 0 |
| Running nose |  |  | 0 | 0 |
| Loss of smell or taste |  |  | 0 | 0 |
| Abdominal pain |  |  | 0 | 0 |
| Loss of appetite |  |  | 0 | 0 |
| Nausea |  |  | 0 | 0 |
| Vomiting |  |  | 0 | 0 |
| Diarrhea |  |  | 0 | 0 |
| Arthralgia |  |  | 0 | 0 |
| Myalgia |  |  | 0 | 0 |
| Itching |  |  | 0 | 0 |
| Skin rash |  |  | 0 | 0 |
| Laboratory abnormalities |  |  |  |  |
| Creatinine |  |  | 0 | 0 |
| BUN |  |  | 0 | 0 |
| Sodium |  |  | 0 | 0 |
| eGFR |  |  | 0 | 0 |
| Potassium |  |  | 0 | 0 |
| ALT/SGPT |  |  | 0 | 0 |
| AST/SGOT |  |  | 0 | 0 |
| Total bilirubin |  |  | 0 | 0 |
| Direct bilirubin |  |  | 0 | 0 |
| Alkaline Phosphatase |  |  | 0 | 0 |
| LDH |  |  | 0 | 0 |
| Creatinine phosphokinase (CPK) |  |  | 1* | 2* |
| Anemia |  |  | 0 | 0 |
| Leukocytopenia |  |  | 0 | 0 |
| Neutropenia |  |  | 0 | 0 |
| Thrombocytopenia |  |  | 0 | 0 |

**Patients were also classified as serious adverse events and are detailed in the serious adverse events table (Table: 2). All AEs solicited were reported between days 0-7, day 14 and at day 28.*

### Adverse events (AE) for casirivimab/imdevimab

Supplementary table 1: Summary of adverse events (grade 3 and above) for casirivimab/imdevimab

|  | **All grades** | | **Grade 3-4** | |
| --- | --- | --- | --- | --- |
|  | **casirivimab/imdevimab (n=74)** | **No study drug (n=89)** | **casirivimab/imdevimab (n=74)** | **No study drug (n=89)** |
| Any adverse event (Grade ≥ 3) | 0 | 2 |  |  |
| Serious adverse event reported | 0 | 2 |  |  |
| Symptoms |  |  |  |  |
| Fever |  |  | 0 | 0 |
| Headache |  |  | 0 | 0 |
| Dizziness |  |  | 0 | 0 |
| Blurred vision |  |  | 0 | 0 |
| Fatigue |  |  | 0 | 0 |
| Cough |  |  | 0 | 0 |
| Difficulty breathing |  |  | 0 | 0 |
| Chest pain |  |  | 0 | 0 |
| Running nose |  |  | 0 | 0 |
| Loss of smell or taste |  |  | 0 | 0 |
| Abdominal pain |  |  | 0 | 0 |
| Loss of appetite |  |  | 0 | 0 |
| Nausea |  |  | 0 | 0 |
| Vomiting |  |  | 0 | 0 |
| Diarrhoea |  |  | 0 | 0 |
| Arthralgia |  |  | 0 | 0 |
| Myalgia |  |  | 0 | 0 |
| Itching |  |  | 0 | 0 |
| Skin rash |  |  | 0 | 0 |
| Laboratory abnormalities |  |  |  |  |
| Creatinine |  |  | 0 | 0 |
| BUN |  |  | 0 | 0 |
| Sodium |  |  | 0 | 0 |
| eGFR |  |  | 0 | 0 |
| Potassium |  |  | 0 | 0 |
| ALT/SGPT |  |  | 0 | 0 |
| AST/SGOT |  |  | 0 | 0 |
| Total bilirubin |  |  | 0 | 0 |
| Direct bilirubin |  |  | 0 | 0 |
| Alkaline Phosphatase |  |  | 0 | 0 |
| LDH |  |  | 0 | 0 |
| Creatinine phosphokinase (CPK) |  |  | 0 | 2* |
| Anemia |  |  | 0 | 0 |
| Leukocytopenia |  |  | 0 | 0 |
| Neutropenia |  |  | 0 | 0 |
| Thrombocytopenia |  |  | 0 | 0 |

**Both patients were also classified as serious adverse events and are detailed in the serious adverse events table (Table: 2). All AEs solicited were reported between days 0-7, 14 and at day 28.*

### Serious Adverse Events

Supplementary table 2: Summary of Serious Adverse Events

| **Number** | **Study arm** | **Final diagnosis** | **Relationship to trial drug** | **Resolved** |
| --- | --- | --- | --- | --- |
| 1 | No study drug | Reduction in activities of daily living after COVID-19 infection ^1^ | Not related | Yes |
| 2 | No study drug | COVID-19-related skeletal muscle damage ^2^ | Not related | Yes |
| 3 | No study drug | COVID-19-related skeletal muscle damage ^2^ | Not related | Yes |
| 4 | Remdesivir | COVID-19-related skeletal muscle damage ^2^ | Not related | Yes |

*^1^Participant was impeded in activities of daily living one day post-discharge from the ward (day 8) and was readmitted for further investigation complaining of right-sided chest pain and lethargy. Clinical observations including oxygen saturations, physical examination and electrocardiogram were unremarkable, laboratory investigations including inflammatory markers and D-Dimers were all in the normal range. A SARS-CoV-2 PCR was negative. CT pulmonary angiogram showed no radiological evidence of pulmonary embolus or pneumonitis. None of the patient’s symptoms were classifiable as grade 3 or above, although because of the hospitalization an SAE was reported. The patient’s symptoms quickly resolved and the patient was discharged the following day.*

*^2^Participants had an acute rise in serum phosphokinase (CPK) during admission. Myoglobin was not detected on urinalysis. A diagnosis of COVID-related skeletal muscle damage was made based on symptoms of myalgia and a raised CPK without another identifiable etiology. Nephrology review advised supportive treatment only. Symptoms resolved, the CPK normalized and was within normal limits on follow-up.*

### Virus variant determination

Brazil site – Virus variant determination

***SARS-CoV-2 whole-genome sequencing***

The sequencing was carried out using two different technologies, Illumina (Illumina, USA) and IonTorrent (ThermoFisher Scientific, USA). Only SARS-CoV-2-positive samples with Ct < 30 values for virus targets were considered. Illumina libraries were prepared using the QIAseq FX DNA Library Prep kit® (QIAGEN, Germany) and sequenced on the Illumina MiSeq® platform (Illumina, USA) with a v3 (600 cycles) cartridge, following all manufacturer’s protocols. IonTorrent libraries were prepared using the Ion AmpliSeq SARS-CoV-2 Panel® (ThermoFisher Scientific, USA) and sequenced on the IonTorrent PGM platform® with a 314-chip kit (ThermoFisher Scientific, USA), according to the manufacturer’s recommendations. Three negative controls were used in all sample processing steps (cDNA synthesis, viral genome amplification, and library preparation).

***Viral genome assembly and classifications***

A custom pipeline was used to process the sequencing data. In the first step, quality control was performed with Trimmomatic v0.39. Adapter and primer sequences, short reads (< 50 nucleotides), and low-quality bases (Phred score < 30) were removed. Next, reads were mapped against the SARS-CoV-2 reference genome (GenBank accession: NC_045512) with Bowtie2. Samtools manipulated the mapping files, whilst consensus genome sequences were estimated using the bcftools consensus option. Masking of low-coverage sites was performed with bedtools. The code for the described pipeline can be found on GitHub (https://github.com/filiperomero2/ViralUnity). Depth thresholds differed between sequencing technologies employed. For IonTorrent data, sites with less than 20-fold depth were masked, while for Illumina, the minimum threshold was 10-fold. Sequences <70% genome coverage breadth were removed from downstream analysis. Consensus sequences were classified using the NextClade web application v.1.7.0, or Pangolin tool v.3.1.11.

Thailand site – Virus variant determination

***SARS-CoV-2 whole-genome sequencing***

The sequencing method carried out in this experiment follows the “PCR tiling of SARS-CoV-2 virus with rapid barcoding and Midnight RT PCR Expansion” provided by Oxford Nanopore Technology (Oxford, UK) developed based on a protocol by ARTIC network group^1^. Library preparation process started with reverse transcription, which consists of mixing the purified viral RNA with LunaScript RT SuperMix and incubating the mixtures in a thermal cycler. DNA fragments to be used in the assembly process were amplified by PCR using Midnight primer set (V3) and attached with barcodes from Rapid Barcode Plate (RB96). The mixtures from each sample were pooled together, cleaned with AMPure XP Beads (AXP) and attached with Rapid Adapter F (RAP F). The prepared DNA fragments were then loaded into a primed flow cell (FLO-MIN106) and sequenced on GridION MK1 system.

***Viral genome assembly and classification***

The output sequencing data (.fast5) from MinKNOW software was base-called with Guppy software using the High Accuracy (HAC) model to generate nucleotide sequence data for each fragment (reads) in the fastq format. These base-called data were then processed through the established workflow wf-artic on EPI2ME software to be assembled into consensus sequences. Only reads with average Phred Quality (Q) score above 9 and minimum and maximum length of 250 and 1500 bps were used in the assembly process. The consensus sequences were then classified using the Pangolin tool (4.1.1) and Pangolin dataset (v1.14).

The virus variants over time are shown in Figure S1.

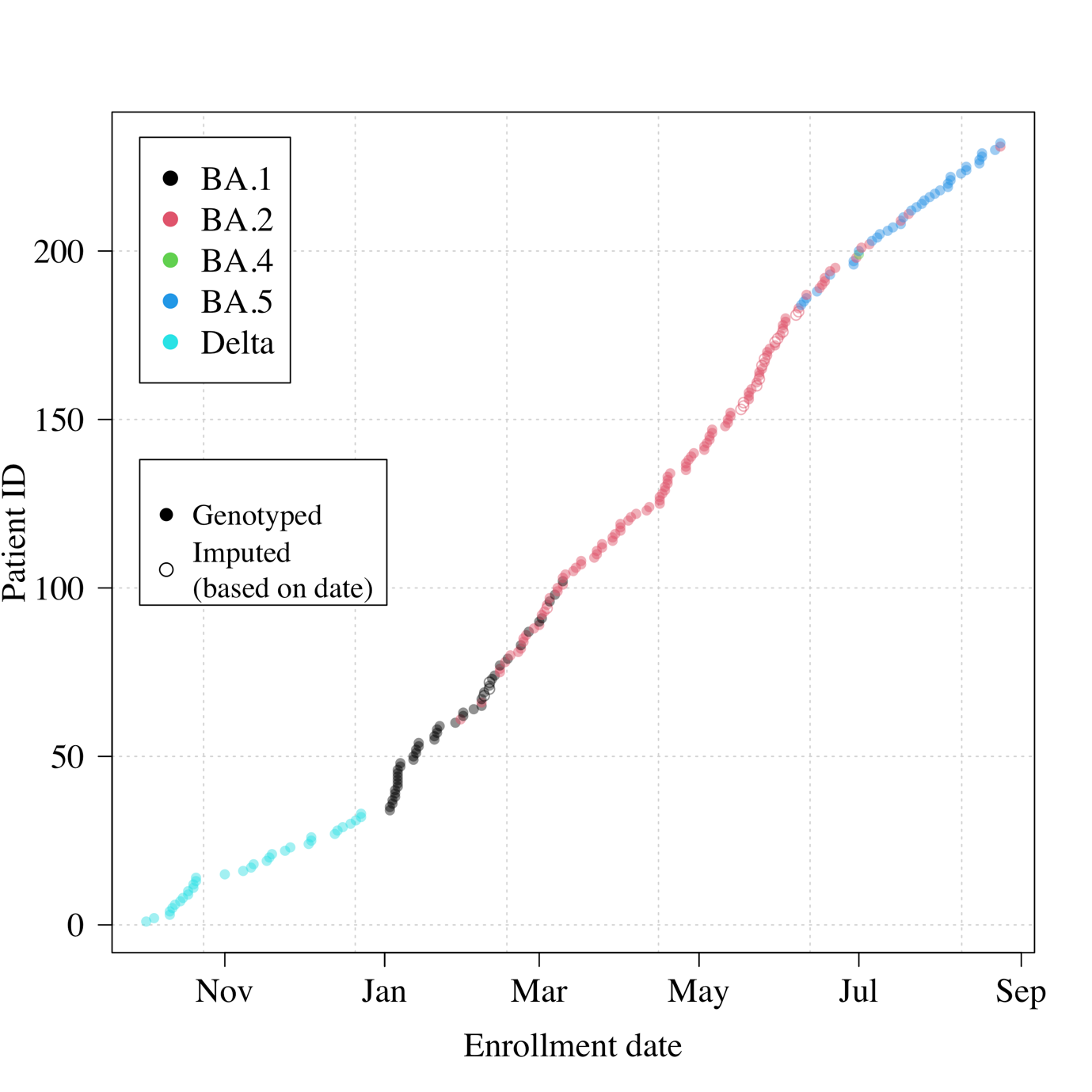

Figure S1 Genotyped variants over time (combined Thai and Brazilian sites). The data from Brazil are imputed based on date.

### Randomization

The randomization sheets were generated by the trial statistician (James Watson).

All new randomization sheets and all updates of existing randomization sheets were done using a pre-written R script which was stored on the randomization Dropbox folder (owner is MORU, under custodianship of the head of MORU IT); this file is a full ‘Professional’ version with history recorded and only the trial statistician and head of IT had access. The file took the following inputs:

- Site codes (e.g. “th001”) for which to generate randomization sheets;
- The set of arms available for randomization in that site;
- The number of arm repeats per block (this is set to the minimum integer such that in each block there is an integer number for each arm);
- The randomization data file from each site (which has the patient numbers for subjects already randomized) named data-XXX.csv (where XXX is the site code), if this does not yet exist a blank .csv (headers only) is generated.

This R script is run every time a new site becomes active and every time the set of available arms changes. The output is a .csv file named rand-XXX.csv (where XXX is the site code). This overwrites the pre-existing file (which can be retrieved from the Dropbox version history). Each time the randomization script is run, this is recorded on a log file*.*

The randomization is done according to the following constraints:

- Blocks of 2*number of available arms;
- Additional ‘fuzziness’ by swapping one patient allocation per block at random (this can be swapped for any of the available arms) – this avoids knowing which arm the last patient per block will receive.

Each time an authorized member of the study team logs onto the web-app this is logged (timestamp and username).

Each time a new patient is randomized this is logged on to the file data-XXX.csv (where XXX is the site code) with the following information:

- Subject number
- Screening number
- Age
- Sex
- Member of study team username
- Timestamp

### Statistical Analysis

The primary analysis consists of fitting Bayesian hierarchical (mixed effects) linear models to the serial log_10_ viral load data up until day 7 (the day 14 data were not used). All models encode residual error as a *t*-distribution with degrees of freedom estimated from the data. The *t*-distribution was chosen for robustness as the residual error is clearly non-Gaussian. The *t*-distribution error model also makes the inferences robust against model mis-specification (particularly for the linear models).^2^ All models include correlated individual random effect terms for both the intercept (baseline viral load) and the slope. All changes to the slope are defined as multiplicative changes on the log scale (i.e. a value of 0 equals no change).

The treatment effect is defined as the proportional change (expressed as a multiplicative term) in the population slope of the daily change in log_10_ viral load. The data are modelled on the log_10_ copies per mL scale, after conversion from Ct values using the standard curve generated from the 12 control concentrations (synthetic samples with known viral densities) from each 96 well plate. The standard curve transformation is done by fitting a linear mixed effects model (random slope and random intercept for each plate) to the control data: regressing the Ct values on the known log viral densities. This borrows information across plates and adjusts for batch effects.

For all models, we adjusted the intercept and slope for the enrolling site (4 sites in total, the reference site is the Hospital of Tropical Diseases which recruited >80% of patients) and for the variant called (Delta is reference: BA.1, BA.2, BA.4, BA.5 are the alternatives). A subset of models also adjusted the slopes and intercepts for:

- Age
- Number of vaccine doses
- Days since symptom onset

All models adjust for human RNase P (proxy for the number of human cells in the sample). This is an independent linear predictor for each viral load measurement. We fitted the models using two sets of prior distributions: weakly informative priors (WIP) and non informative priors (NIP).

***Models fitted***

For each analysis we fitted 5 separate models:

1. Model 1 is linear with RNase P adjustment; adjustment for site & variant; WIP. **This is the main model used to report treatment effects.**
2. Model 2 is non-linear; RNase P adjustment; adjustment for site & variant; WIP
3. Model 3 is linear with RNase P adjustment; adjustment for site & variant; non-informative priors (NIP)
4. Model 4 is non-linear with RNase P adjustment; adjustment for site & variant; NIP
5. Model 5 is linear with RNase P adjustment; full covariate adjustment; WIP

Model 1 was used for all stopping decisions. All models have RNase P adjustment and are all combinations of linear & non-linear models, with or without full covariate adjustment; and with either weakly informative priors or non-informative priors. We compared model fits using the *loo* (approximate leave-one-out cross validation) package. The statistical analysis plan provides a detailed overview of the model structures.

***Analyses***

Adaptive platform trials can suffer from temporal confounding. The main analyses for the remdesivir and casirivimab/imdevimab arms used concurrent controls only (i.e. patients in the no study drug arm who could have been randomised to the active arm). For remdesivir, this included all controls in Thailand and Brazil enrolled up until the 10^th^ June 2022. For casirivimab/imdevimab this included all controls in the Thai sites only (the mAbs were not available in Brazil) up until the 24^th^ August 2022. In addition, we did two extra analyses, one with all concurrent remdesivir, casirivimab/imdevimab, and no study drug patients in Thailand up until the 10^th^ June (all concurrent), and one with all patients.

The virus variant subgroup analyses were fitted with an interaction term between casirivimab/imdevimab and the virus subgrouping of interest (three subgroupings were analysed in total). The same WIP prior (Normal[0,0.5]) was used for the subgroup interaction term as for the main effect (note this pulls effects in small subgroups towards the overall effect).

The analyses were structured as follows (for each analysis 5 models as specified above were fitted):

1. Remdesivir versus no study drug (all patients up until 10^th^ June; analysis data set contains 131 patients and 2,356 viral load measurements).
2. Casirivimab/imdevimab versus no study drug (all Thai patients up until 24^th^ August; analysis data set contains 158 patients and 2,842 viral load measurements).
3. Contemporaneous remdesivir and casirivimab/imdevimab (all Thai patients up until 10^th^ June) with subgroup effects of the casirivimab/imdevimab by WHO major greek lineage (Delta vs Omicron; analysis data set contains 169 patients and 3,040 viral load measurements).
4. Same as 2 (casirivimab/imdevimab versus no study drug) with an interaction term for Delta vs Omicron virus variants (main pre-specified subgroup analysis; analysis data set contains 158 patients and 2,842 viral load measurements).
5. Same as 2 (casirivimab/imdevimab versus no study drug) with an interaction term for Delta vs Omicron G446S vs Omicron not G446S virus variants (posthoc subgroup analysis; analysis data set contains 158 patients and 2,842 viral load measurements).
6. Same as 2 (casirivimab/imdevimab versus no study drug) with an interaction term for Delta vs BA.1 vs BA.2 vs BA.5 (posthoc subgroup analysis, this excludes BA.4 as there was only one patient; analysis data set contains 157 patients and 2,824 viral load measurements).
7. All data (adjusting for site and variant; analysis data set contains 232 patients and 4172 viral load measurements).

Thus a total of 35 models were fitted to the data. The main text reports effect estimates from analyses 1 and 2. However as the full analysis (analysis 7) gave nearly identical treatment effect estimates and had the largest total sample size, we use it to report covariate effects (e.g. differences in clearance between virus variants).

***Code and analysis plan***

All data, models and analytical output are on the linked github repository: <https://github.com/jwatowatson/PLATCOV-Remdesivir-Regeneron>.

This includes all data used in the analysis for full reproducibility of the results. The main RMarkdown file (Full_Analysis.Rmd) goes through all data manipulation and model fitting. The Statistical Analysis Plan used (version 2.2) is also provided in this repository.

### Stopping rules

The stopping rules were determined using a simulation approach, based on previously modelled serial viral load data (3), such that approximately 50 patients are needed to demonstrate increases in the rate of viral clearance of ~50%, with control of both type 1 and type 2 errors at 10%. The prespecified decision criteria for stopping a treatment arm were either a model-based probability of <0.1 that the intervention did not accelerate viral clearance by >12.5% relative to no study drug (futility), or >0.9 that it did (success). The first interim analysis (n=50) was prespecified as unblinded in order to review the methodology and stopping rules. Following this, the stopping threshold was increased from 5% to 12.5% because the treatment effect in the positive control arm (casirivimab/imdevimab) in SARS-CoV-2 Delta variant infections was substantially larger than expected. Thereafter trial investigators were blinded.

### Supplementary Results

#### Analysis 1: remdesivir versus no study drug

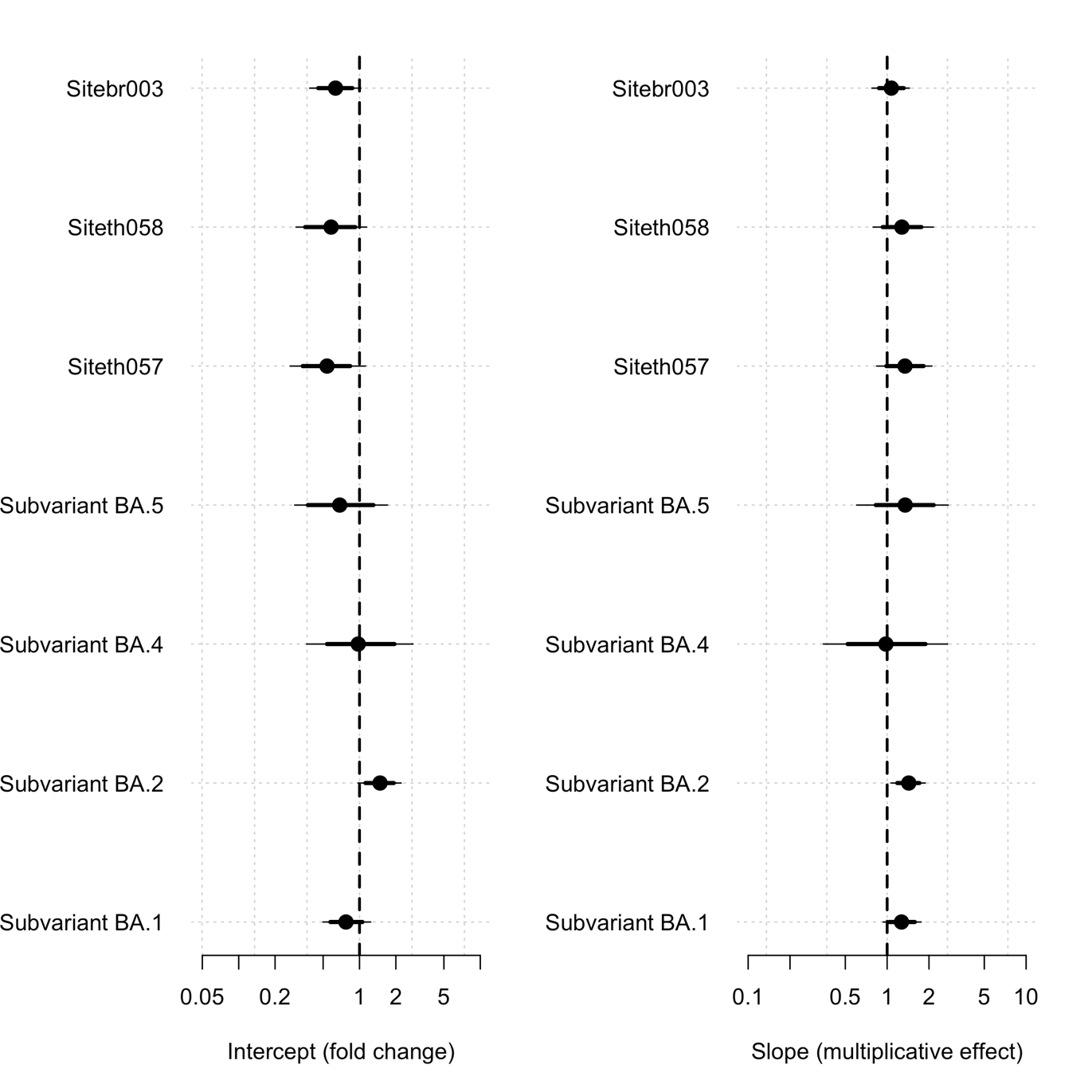

Figure S1: Covariate effects (mean estimate with 80% and 95% credible intervals shown by the thick and thin lines respectively) estimated for the main analytical linear model (model 1 see Statistical Analysis) in the remdesivir versus no study drug analysis . For the site covariates (br003: Brazil; th058: Bangplee; th057: Vajira) the estimate shows the fold change in intercept and slow relative to th001 (FTM). For the variants it is relative to Delta. Dashed vertical line shows value of 1 (no change).

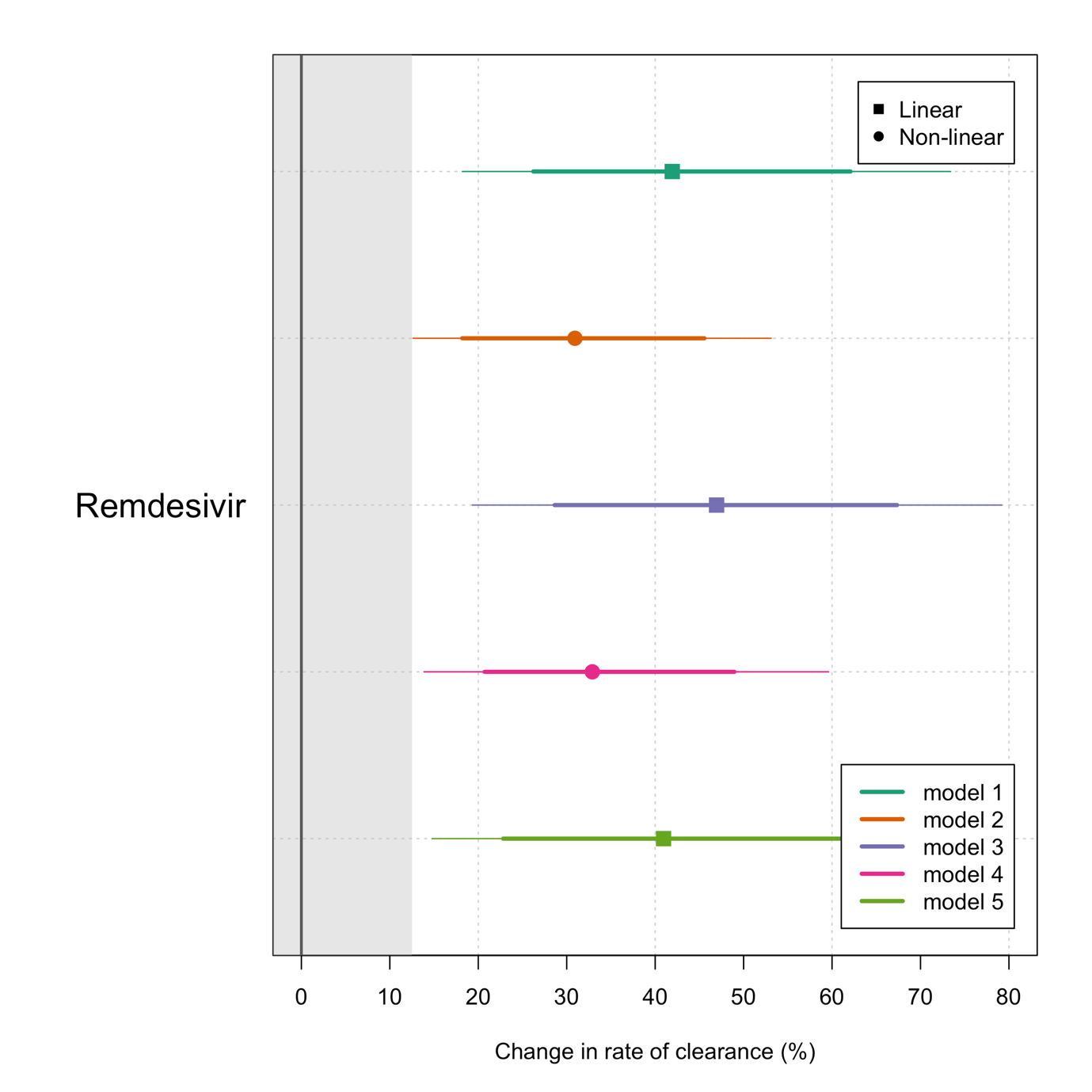

Figure S2 Treatment effects (mean estimate with 80% and 95% credible intervals shown by the thick and thin lines respectively) for remdesivir versus no study drug under the 5 analytical models (see Statistical Analysis).

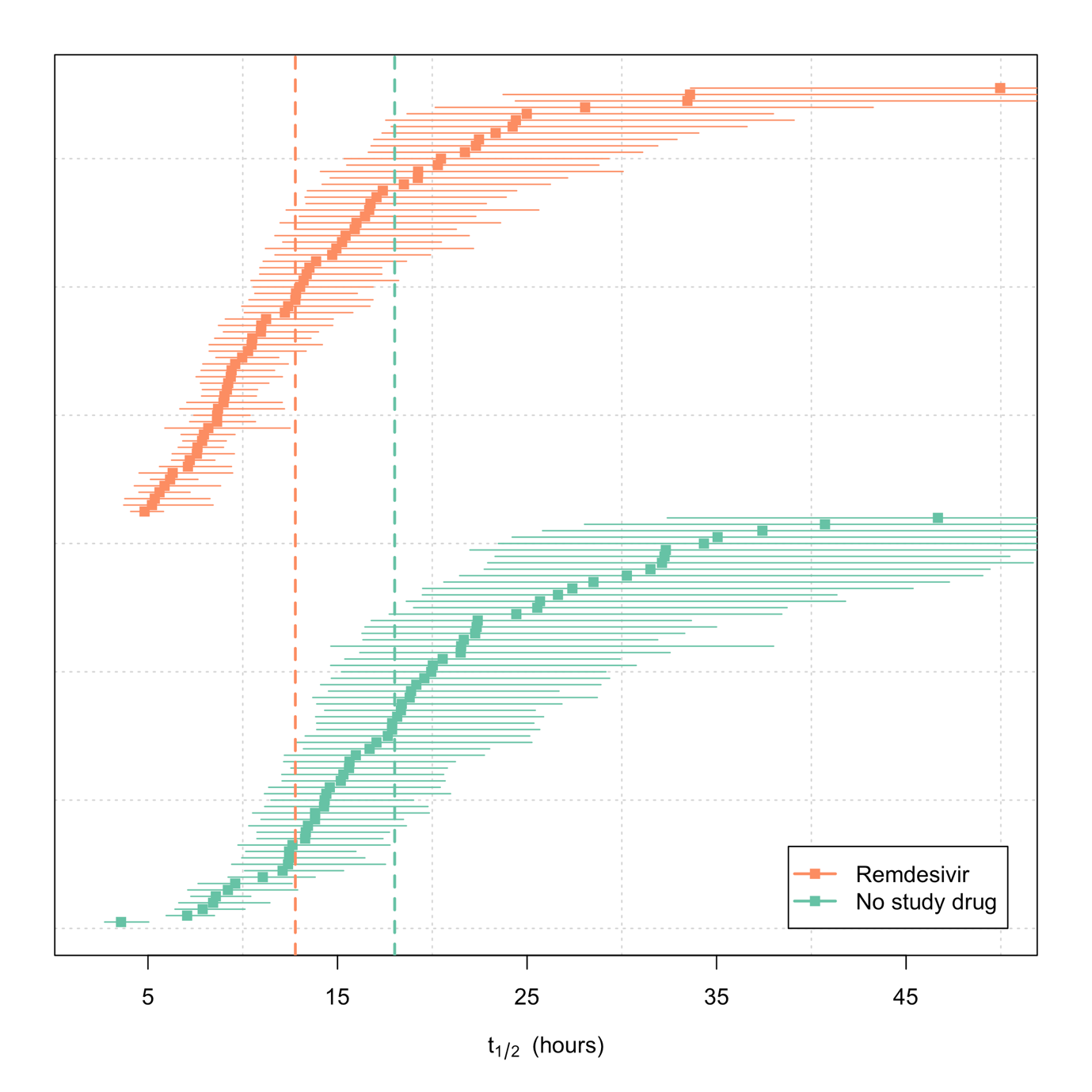

Figure S3 Distribution of clearance half lives in the remdesivir (orange) and no study drug arms (green). The mean estimate is show by the squares, the 95% credible interals are shown by the think lines. We truncate the plot at 50 hours as the uncertainty is highly skewed to the right. The vertical dashed lines show the median (of the mean estimates) half-lives for each arm.

#### Analysis 2: casirivimab/imdevimab versus no study drug (all variants combined)

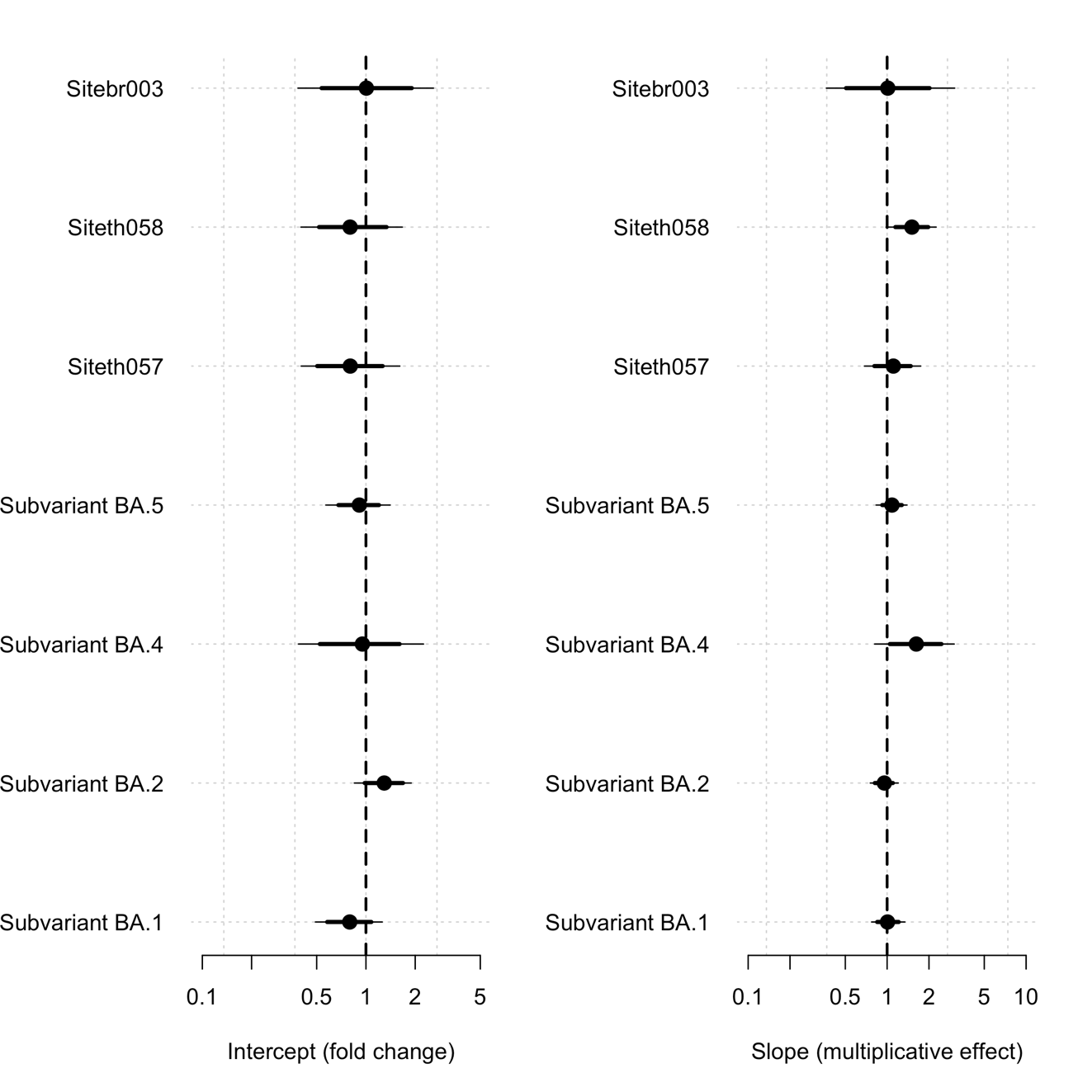

Figure S4: Covariate effects (mean estimate with 80% and 95% credible intervals shown by the thick and thin lines respectively) estimated for the main analytical linear model (model 1 see Statistical Analysis) in the casirivimab/imdevimab versus no study drug analysis. For the site covariates (br003: Brazil; 058: Bangplee; 057: Vajira) the estimate shows the fold change in intercept and slow relative to th001 (FTM). For the variants it is relative to Delta. Dashed vertical line shows the value of 1 (no change).

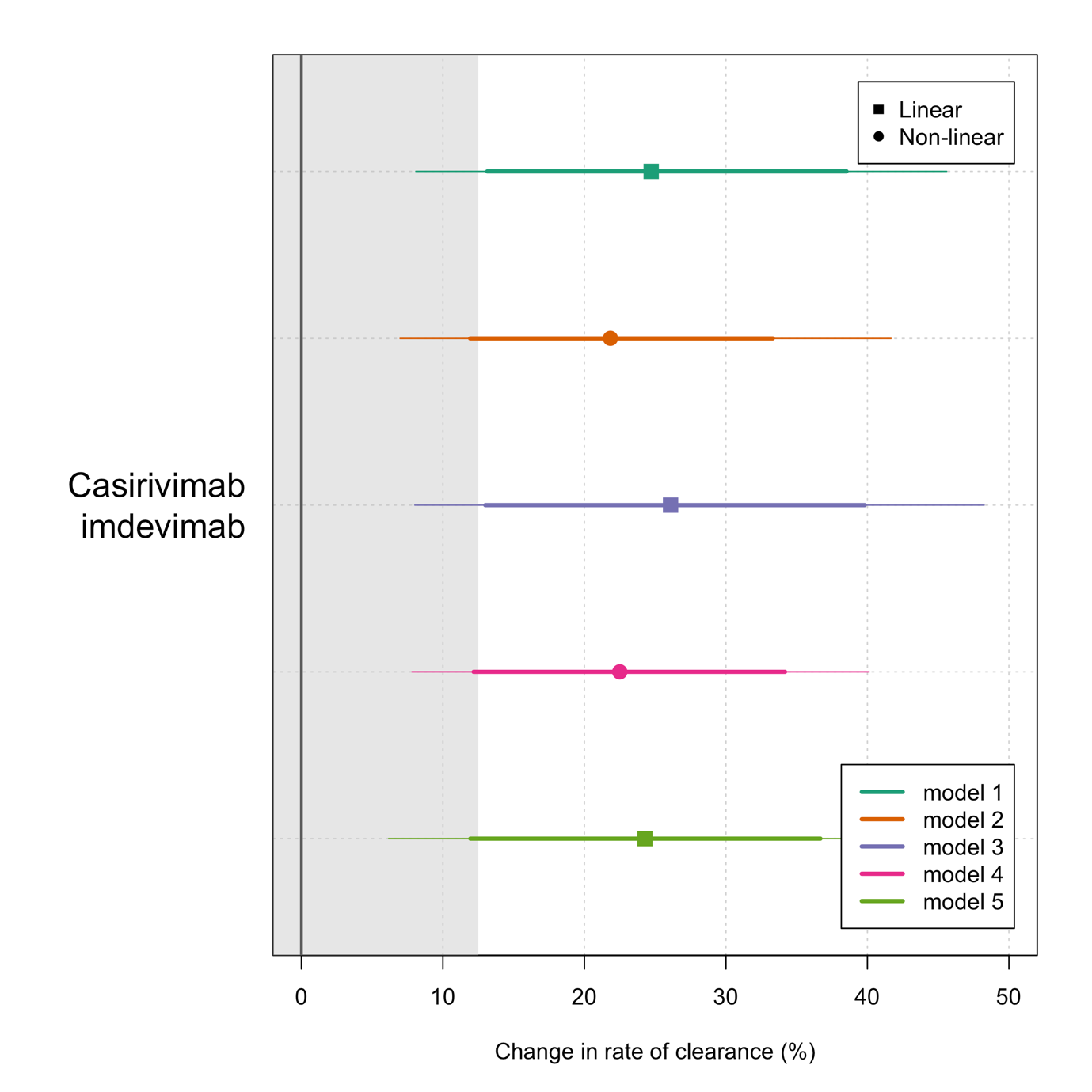

Figure S5 Treatment effects (mean estimate with 80% and 95% credible intervals shown by the thick and thin lines respectively) for casirivimab/imdevimab versus no study drug under the 5 analytical models- see Statistical Analysis (analysis assuming the same effect for all variants).

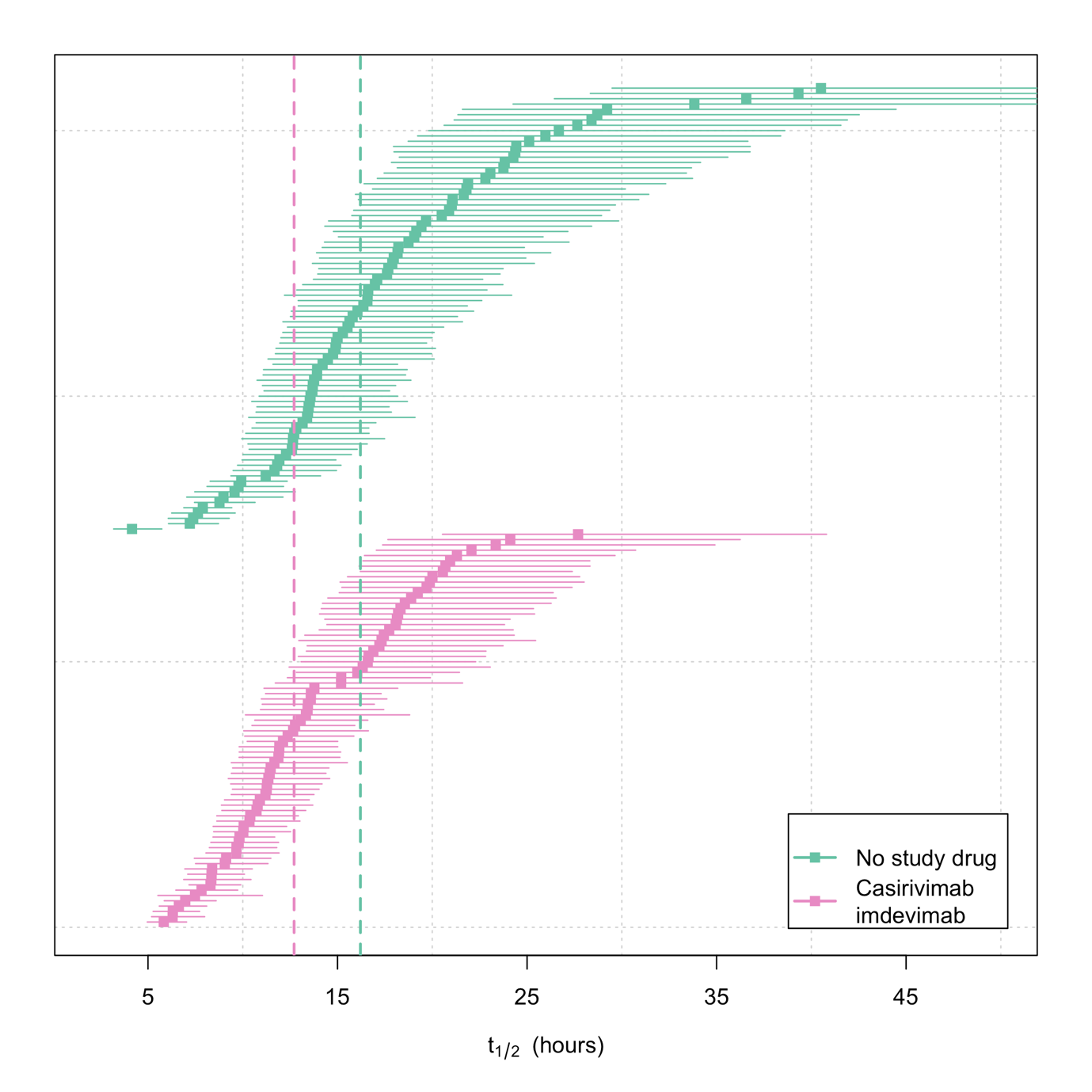

Figure S6 Distribution of clearance half lives in the casirivimab/imdevimab (purple) and no study drug arms (green). The mean estimate is show by the squares, the 95% credible interals are shown by the think lines. We truncate the plot at 50 hours as the uncertainty is highly skewed to the right. The vertical dashed lines show the median (of the individual mean estimates) half-lives for each arm.

#### Analysis 3: casirivimab/imdevimab and remdesivir versus no study drug (contemporaneous patients in Thailand)

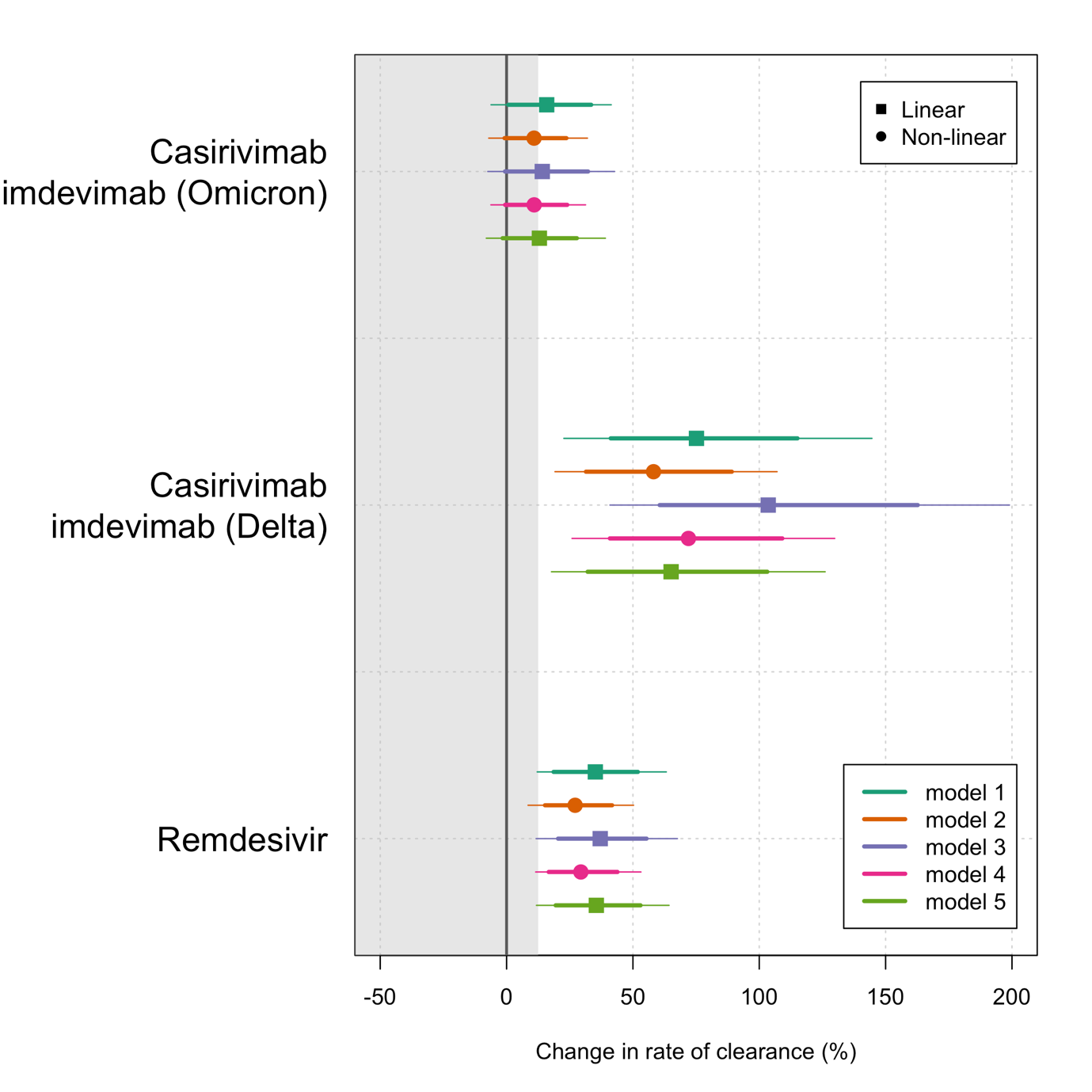

Figure S7 Treatment effects (mean estimate with 80% and 95% credible intervals shown by the thick and thin lines respectively) for casirivimab/imdevimab and remdesivir versus no study drug under the 5 analytical models- see Statistical Analysis (analysis allowing for varying effects by major Greek lineage). This includes Thai patients only up until 10^th^ June 2022 (contemporaneous patients).

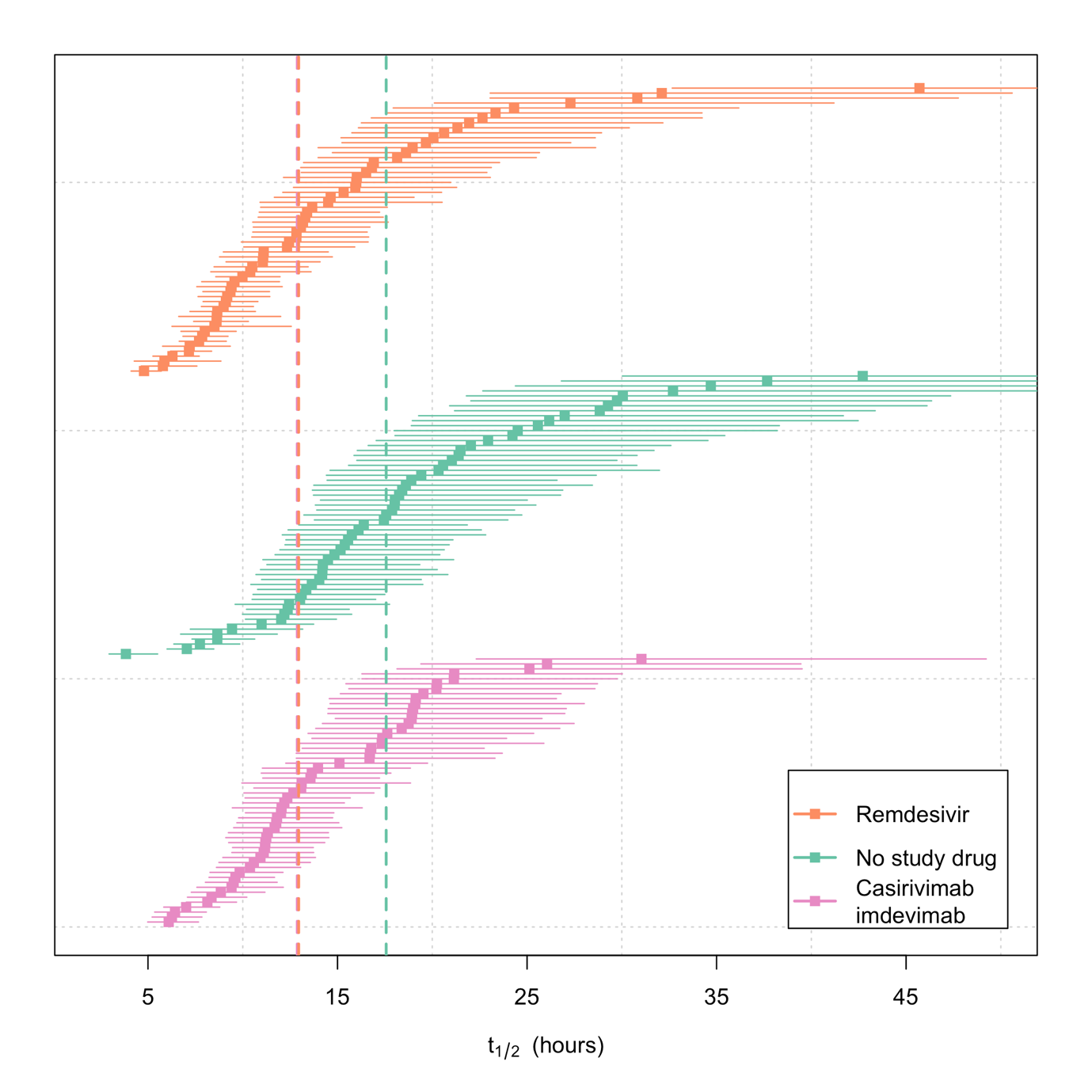

Figure S8 Distribution of clearance half lives in the casirivimab/imdevimab (purple), remdesivir (orange) and no study drug arms (green). The mean estimate is show by the squares, the 95% credible interals are shown by the thick lines. We truncate the plot at 50 hours as the uncertainty is highly skewed to the right. The vertical dashed lines show the median (of the individual mean estimates) half-lives for each arm (nearly identical for remdesivir and casirivimab/imdevimab).

#### Analysis 4: casirivimab/imdevimab versus no study drug with subgroup effects by WHO major Greek lineage (pre-specified subgroup analysis)

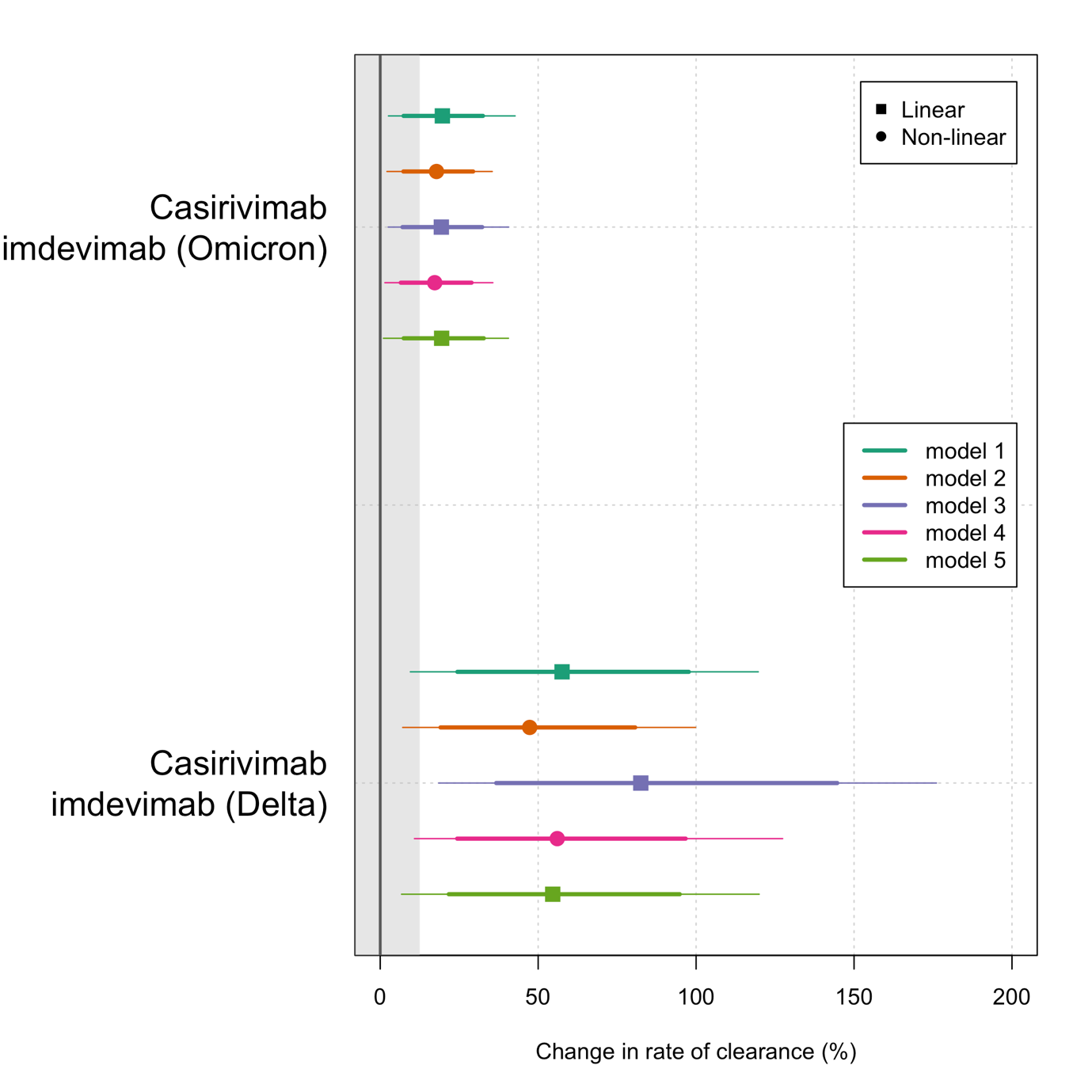

Figure S9 Treatment effects (mean estimate with 80% and 95% credible intervals shown by the thick and thin lines respectively) for casirivimab/imdevimab versus no study drug under the 5 analytical models- see Statistical Analysis (analysis allowing for varying effects by major Greek lineage: the reference effect is in the Delta variant).

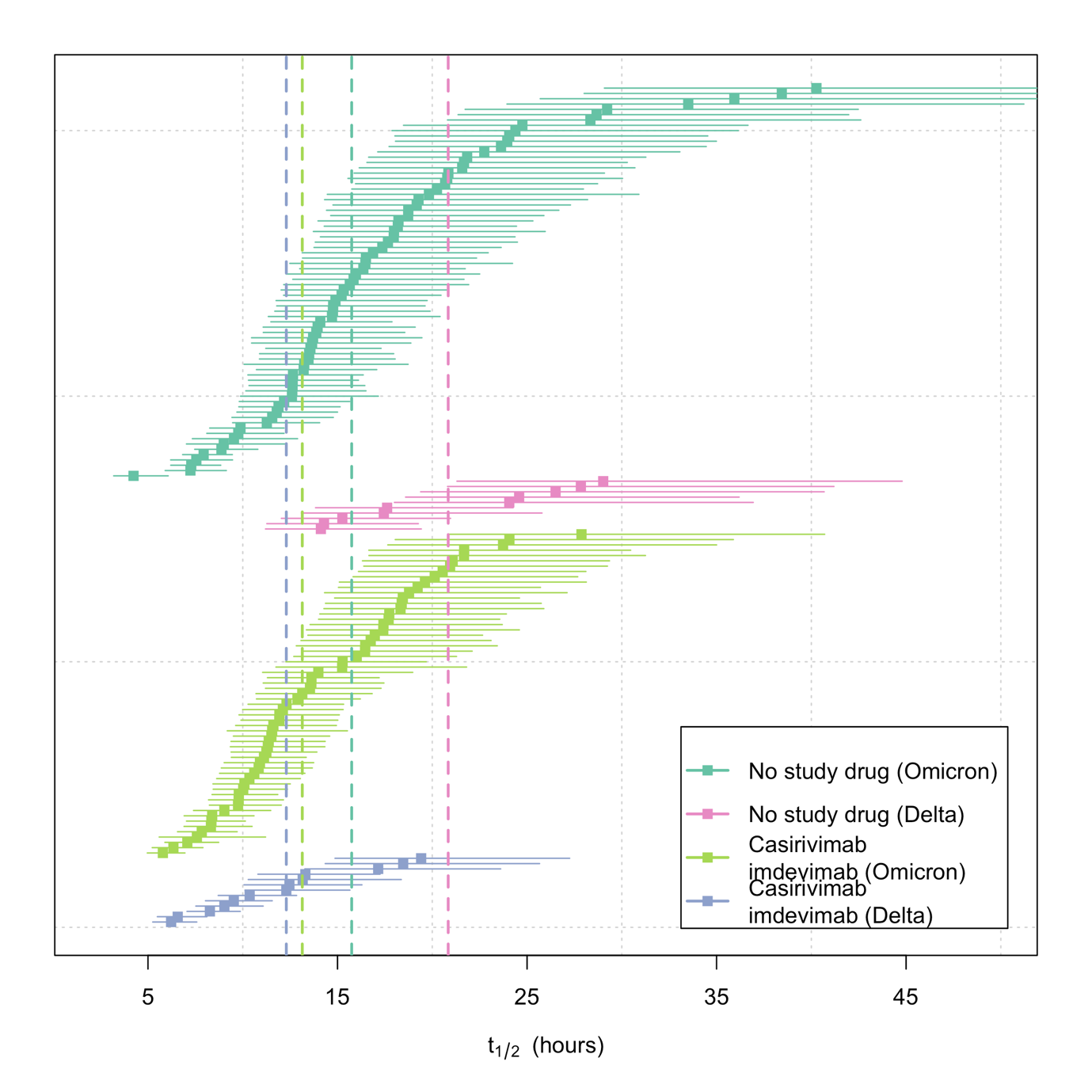

Figure S10. Distribution of clearance half lives in the casirivimab/imdevimab (purple: Delta; light green: Omicron) and no study drug arms (pink: Delta; green: Omicron). The mean estimate is show by the squares, the 95% credible interals are shown by the think lines. We truncate the plot at 50 hours as the uncertainty is highly skewed to the right. The vertical dashed lines show the median (of the individual mean estimates) half-lives for each arm by major Greek lineage.

#### Analysis 5: casirivimab/imdevimab versus no study drug with subgroup effects by G446S mutation (*post hoc* subgroup analysis)

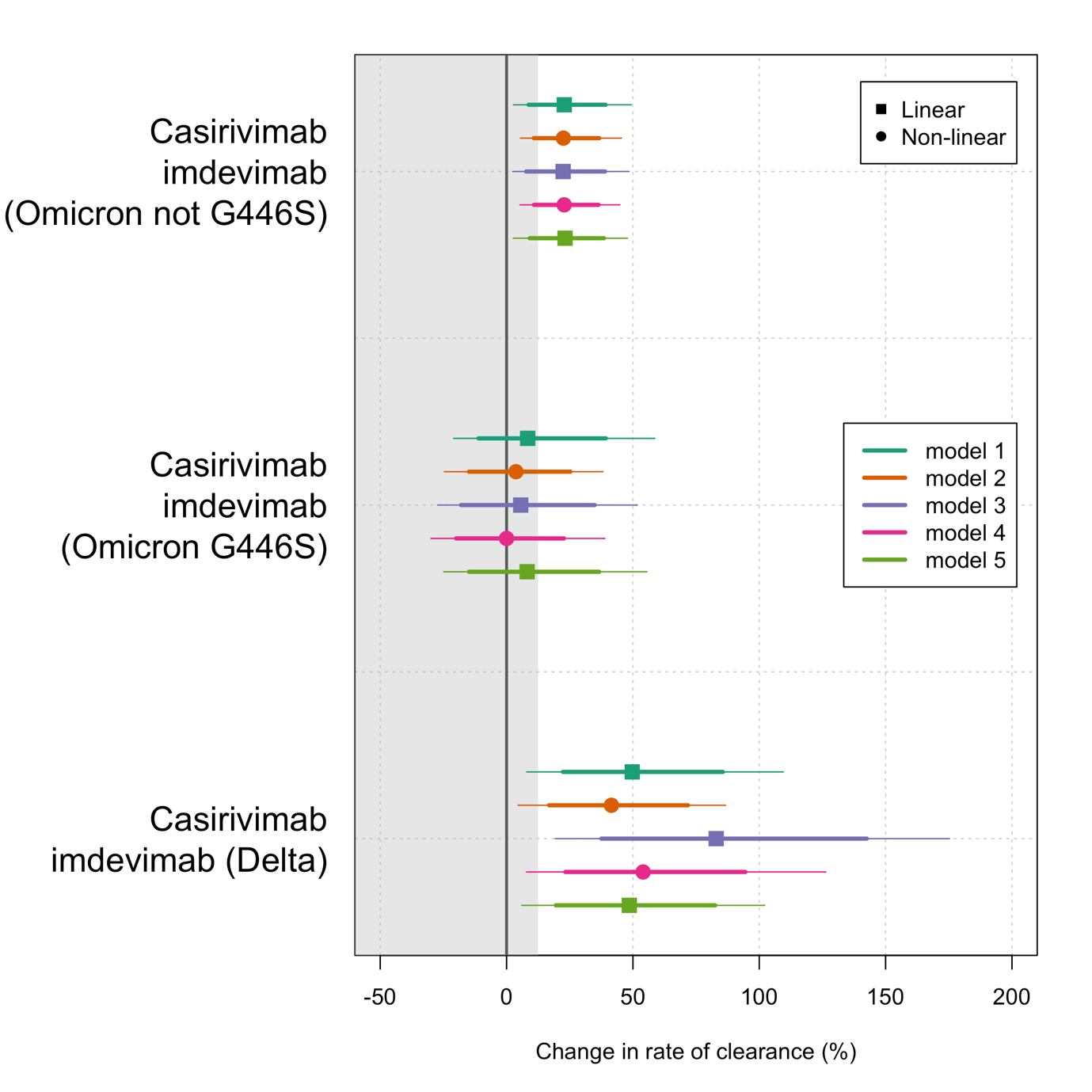

Figure S11 Treatment effects (mean estimate with 80% and 95% credible intervals shown by the thick and thin lines respectively) for casirivimab/imdevimab versus no study drug under the 5 analytical models- see Statistical Analysis (analysis allowing for varying effects by major Greek lineage and by the G446S mutation: the reference effect is in the Delta variant).

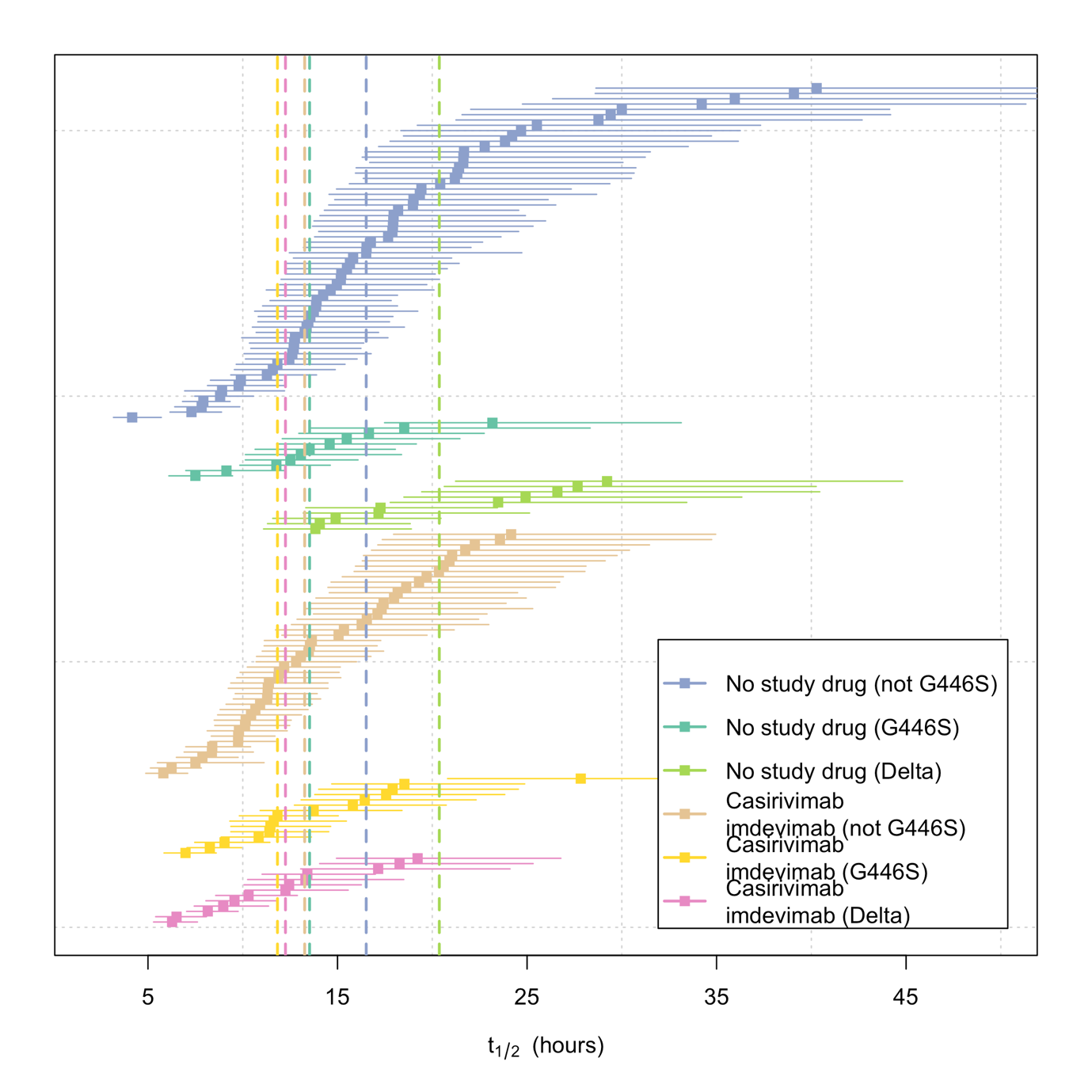

Figure S12 Distribution of clearance half lives in the casirivimab/imdevimab (pink: Delta; light yellow: Omicron G446S; brown: Omicron not G446S) and no study drug arms (light green: Delta; dark green: Omicron G446S; purple: omicron not G446S). The mean estimate is show by the squares, the 95% credible interals are shown by the think lines. We truncate the plot at 50 hours as the uncertainty is highly skewed to the right. The vertical dashed lines show the median (of the individual mean estimates) half-lives for each arm by each grouping.

#### Analysis 6: casirivimab/imdevimab versus no study drug with subgroup effects by major variant sub-lineage (*post hoc* subgroup analysis)

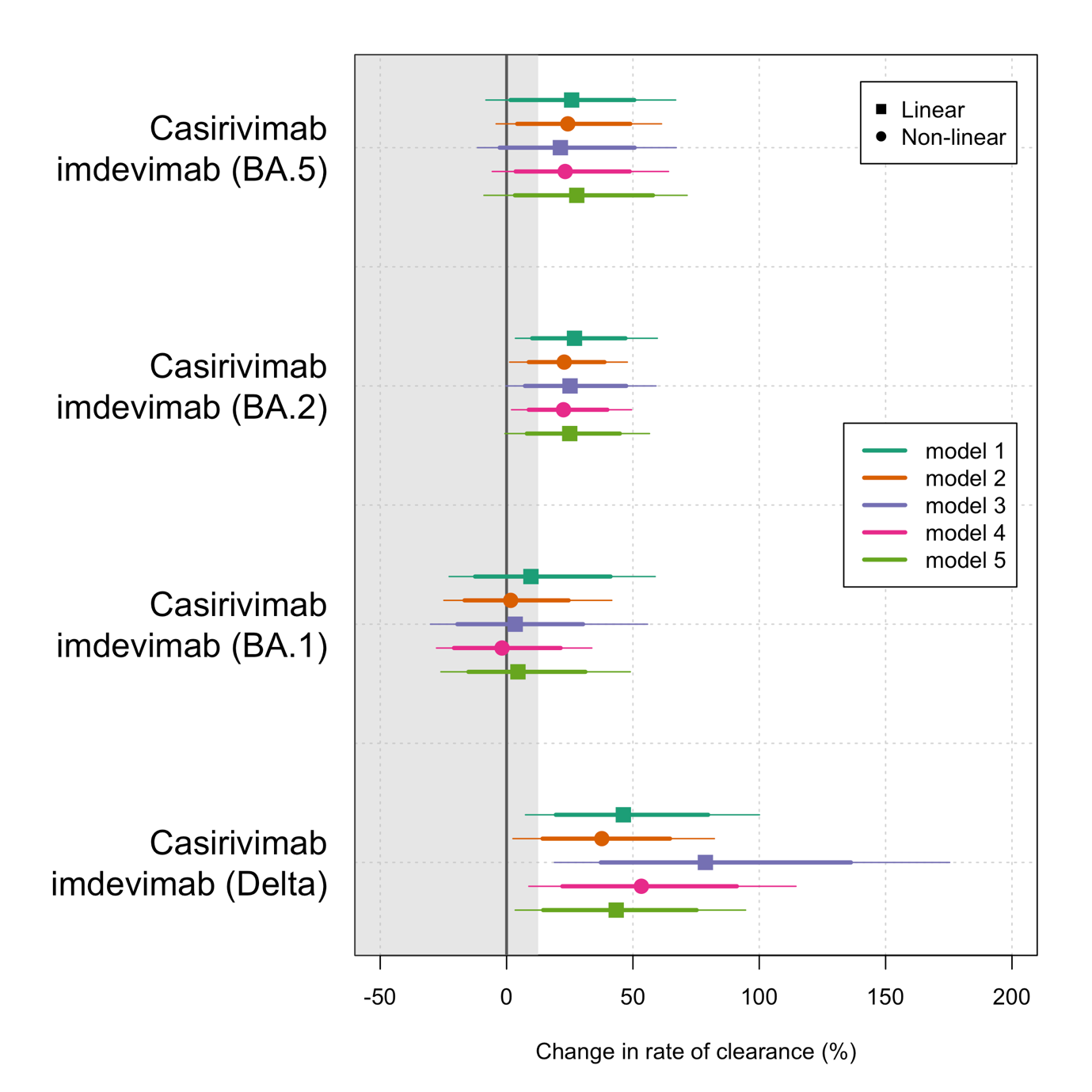

Figure S13. Treatment effects (mean estimate with 80% and 95% credible intervals shown by the thick and thin lines respectively) for casirivimab/imdevimab versus no study drug under the 5 analytical models- see Statistical Analysis (analysis allowing for varying effects by major sub-lineages: the reference effect is in the Delta variant).

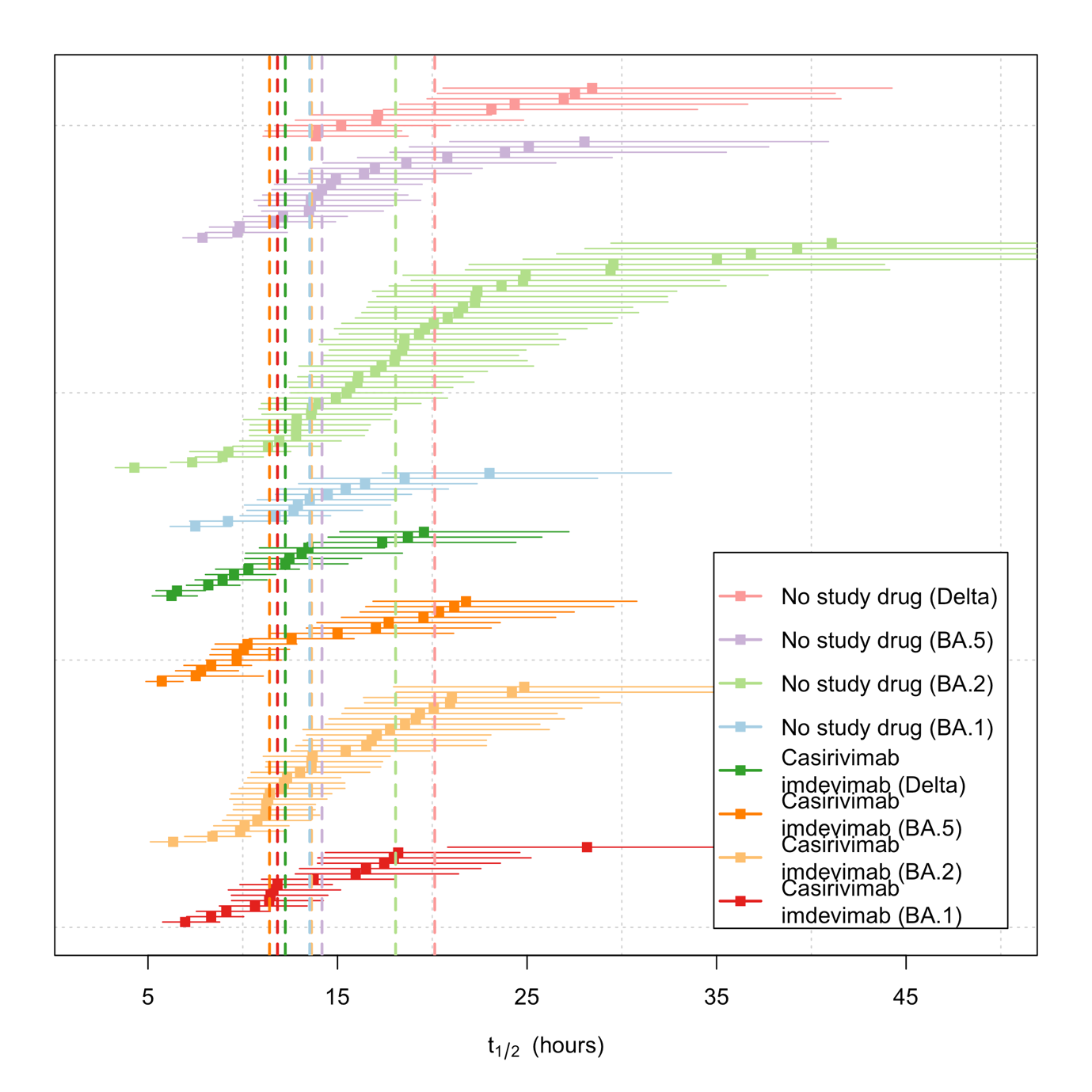

Figure S14. Distribution of clearance half lives in the casirivimab/imdevimab (pink: Delta; light purple: BA.5; light green:BA.2; light blue: BA.1) and no study drug arms (dark green: Delta; orange: BA.5; light orange: BA.2; red: BA.1). The mean estimate is show by the squares, the 95% credible interals are shown by the think lines. We truncate the plot at 50 hours as the uncertainty is highly skewed to the right. The vertical dashed lines show the median (of the individual mean estimates) half-lives for each arm by each grouping.

#### Analysis 7: all data with subgrouping of casirivimab/imdevimab by major Greek lineage

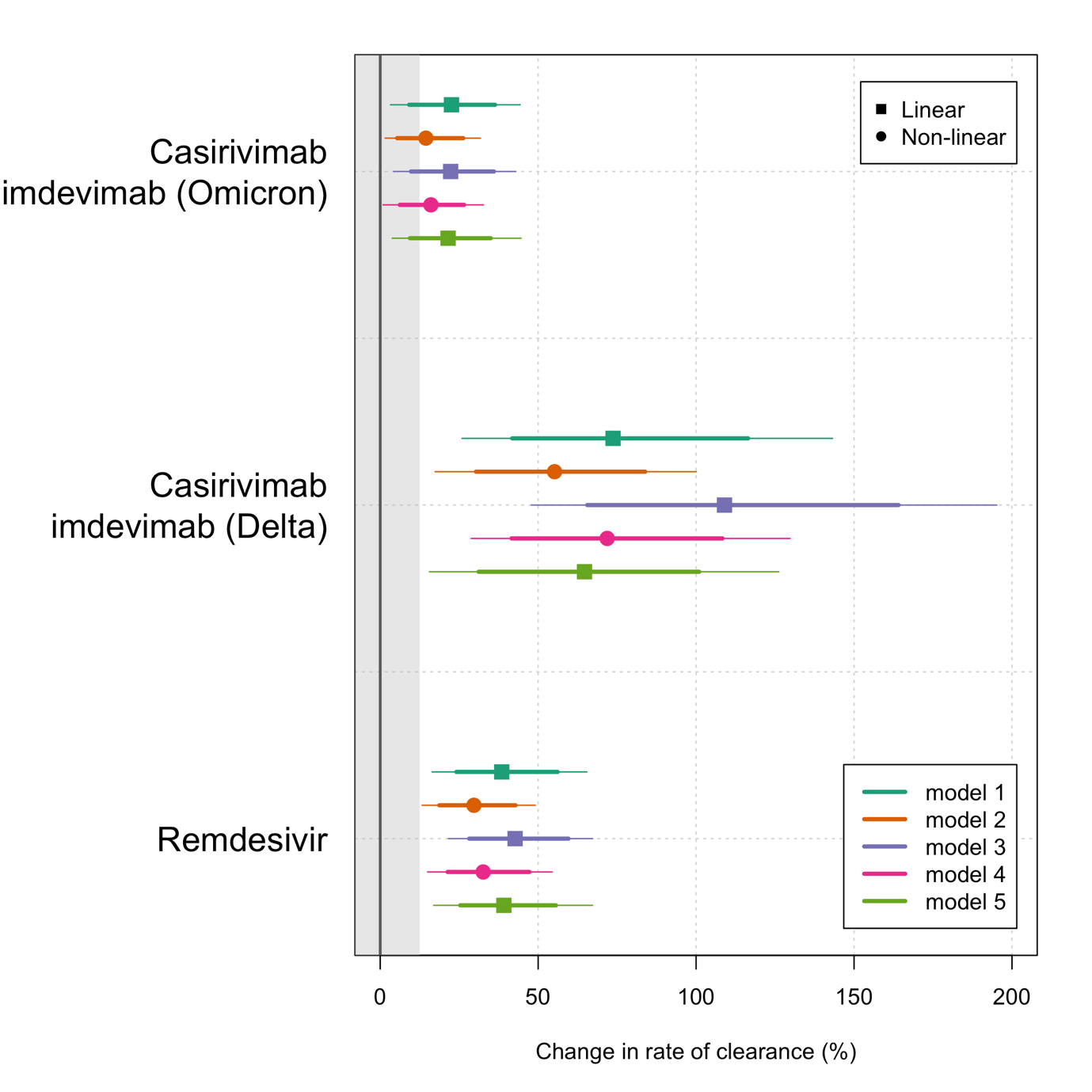

Figure S15. Treatment effects (mean estimate with 80% and 95% credible intervals shown by the thick and thin lines respectively) for casirivimab/imdevimab and remdesivir versus no study drug under the 5 analytical models- see Statistical Analysis (analysis allowing for varying effects of casirivimab/imdevimab by major Greek lineages: the reference effect is in the Delta variant). This analysis includes all patients in the two mITT populations.

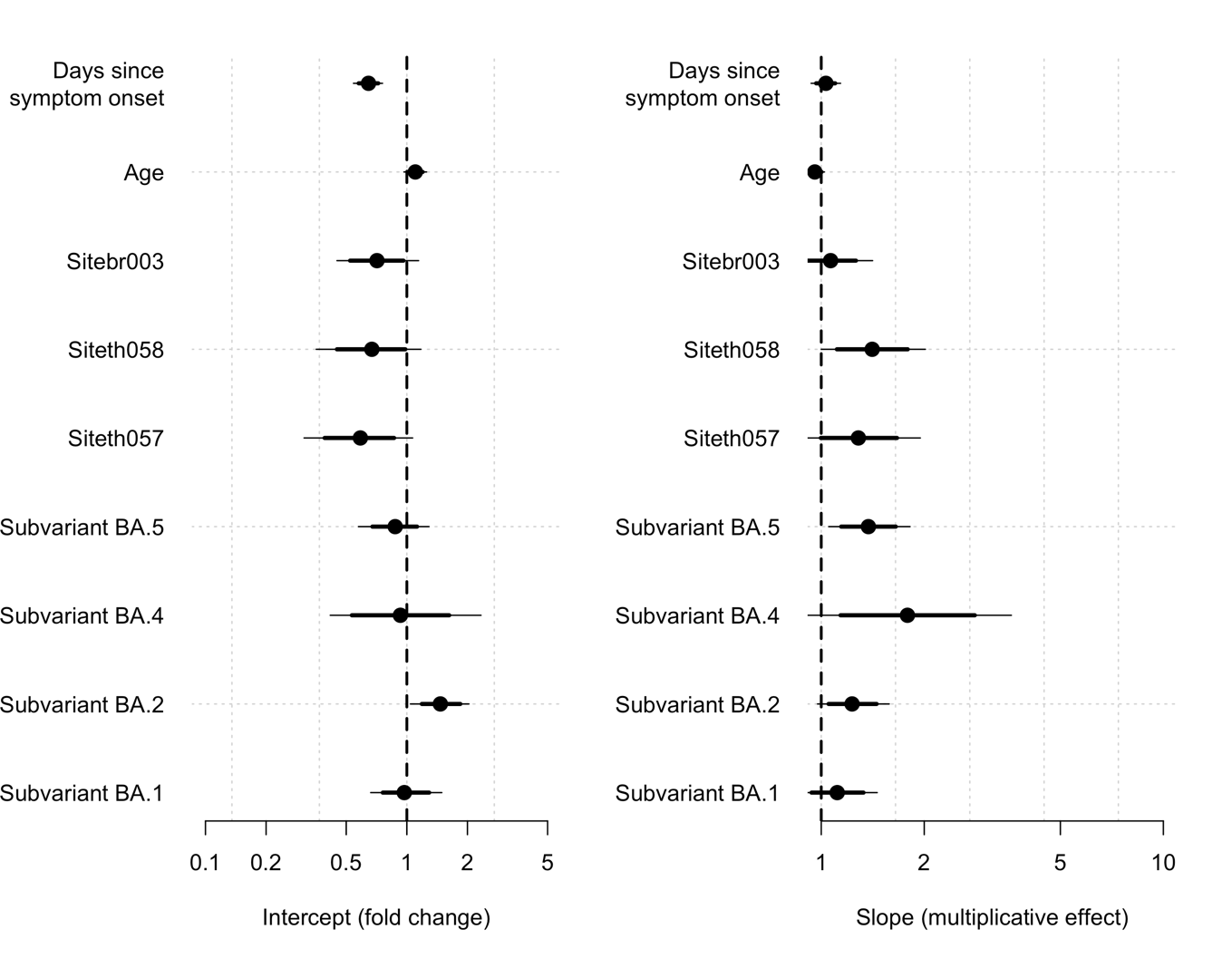

Figure S16. Covariate effects (mean estimate with 80% and 95% credible intervals shown by the thick and thin lines respectively) estimated under linear model with full covariate adjustment (model 5- see Statistical Analysis) analysing all patients in both mITT populations. For the site covariates (br003: Brazil; 058: Bangplee; 057: Vajira) the estimate shows the fold change in intercept and slow relative to th001 (FTM). For the variants the effect is relative to Delta. Age: change for each standard deviation change in age (years); days since symptom onset: change for each additional day since symptom onset.
