## Supplementary material for "Clinical antiviral efficacy of remdesivir and casirivimab/imdevimab against the SARS-CoV-2 Delta and Omicron variants": Latest Statistical Analysis Plan

### Finding Treatments for COVID-19: A Trial of Antiviral Pharmacodynamics in Early Symptomatic COVID-19 (PLATCOV): Statistical Analysis Plan

February 28, 2022

Registered at clinicaltrials.gov number **NCT05041907**

**Version 1**; refers to Master Protocol version V0.1

**Written by:**
Dr James Watson (Statistician)
Signature: Date: 28 February 2022

**Reviewed by and approved by:**

Prof Sir Nicholas White (Co-PI)
Signature: Date: 28 February 2022

Dr William Schilling (Co-PI)
Signature: Date: 28 February 2022

### 1 Trial Overview

An effective, well tolerated, safe, affordable and generally available treatment of COVID-19 that prevented progression to severe disease would be of enormous global health benefit. There are many potential antiviral therapeutics for COVID-19, mainly consisting of repurposed small molecule drugs. At time of writing, however, there is strong evidence for clinical benefit only for the monoclonal antibodies (notably the Regeneron cocktail REGN-CoV-2 [1]) and the small molecule drugs molnupiravir [2] and nirmatrelvir [3]. The clinical benefits of these interventions were determined from large phase 3 studies (>1000 patients) in high-risk individuals enrolled shortly after the onset of a symptomatic SARS-CoV-2 infection. The primary endpoint was most commonly either hospitalisation or death from COVID-19 (usually less than 10% event rate). As vaccine availability becomes widespread in addition to high levels of natural immunity from infection, conducting these types of clinical trials will become more and more difficult as the event rates for the primary endpoints become increasingly low. This calls for improved study designs which allow for the assessment of antiviral interventions in an efficient manner (hundreds rather than thousands of patients). Currently there is no consensus pharmacometric methodology to determine which antivirals should be prioritised for large phase 3 and 4 evaluation, and how to compare current candidates [4].

PLATCOV is a multi-country platform adaptive trial which will develop and validate a methodology for the quantitative assessment of antiviral effects in low-risk patients with high viral burdens and uncomplicated COVID-19. We choose patients with a low risk of progression as this justifies a no treatment arm; we target patients with high viral loads as this is a subgroup in whom antiviral effects can be detected more easily.

This statistical analysis plan (SAP) covers analyses for both the interim and the final reports. We developed the SAP following the Guidelines for the Content of Statistical Analysis Plans in Clinical Trials [5]. It includes pre-specified decision rules for continuing or stopping individual trial arms based on effectiveness or futility.

#### 1.1 Main research questions

Each clinical site in this platform trial will test multiple interventions simultaneously (depending on local regulations and drug availability). The control arm consists of no intervention other than antipyretics and will be compared to intervention arms of three distinct types:

- Newly available or repurposed antiviral drugs (currently randomizing patients to remdesivir, ivermectin, favipiravir, nitazoxanide)
- Positive control (currently REGN-CoV2)
- Novel small molecule drugs (none currently)

There are distinct primary objectives for each intervention type. For the newly available and repurposed drugs, we want to characterise their antiviral activity and compare them to the current gold-standard treatment (currently Regeneron monoclonal antibody cocktail). Many of these drugs are already used and recommended in some countries. Showing that they do not have clinically significant antiviral activity is as important as showing that they do.

For the positive control (currently Regeneron), there is very good evidence from large phase 3 studies that monoclonal antibodies reduce viral load in COVID-19 and reduce hospitalisation in high-risk individuals [1]. However, monoclonal antibodies are vulnerable to viral escape mutations. Recent in vitro data suggest that Regeneron does not have clinically significant neutralising activity against the now dominant Omicron variant [6]. Tracking the performance of the available monoclonals over time is important to characterise the impact and inform the therapeutics of mutant SARS-CoV-2 strains. Monoclonal antibodies are expensive and cannot be produced at large scale currently, but this may change in the near future. These drugs may not be available early in the study in all sites, and will be included depending on local availability and regulatory approval. For any new small molecule drugs in development that pass phase 1 testing, we want to rapidly demonstrate antiviral activity (they will have a stronger rationale, i.e. a priori are more likely to be effective) compared to available drugs. In this case, showing superiority or non-inferiority compared to a gold-standard becomes the key objective. The secondary objectives of this study are:

- To characterise the determinants of viral clearance in early symptomatic SARS-CoV-2 (e.g. estimate the contribution of age, baseline serology, virus genotype, and prior vaccination);
- To determine optimal dosing regimens for drugs shown to have considerable antiviral activity (for interventions of type A and C that have >0.9 probability of increasing viral clearance rate >5%: we will conduct a pharmacokinetic-pharmacodynamic sub-study);
- To compare the antiviral activity of active interventions against current standard of care monoclonal antibodies.

The tertiary objective of this study is to characterise the relationship between viral clearance and the risk of subsequent hospitalisation or death by day 28. However, the event rate in the enrolled population is likely to be extremely low (we hope 0%) so it is very unlikely that we will be able to demonstrate any link between viral clearance and progression to severe COVID-19.

### 2 Study Methods

#### 2.1 Trial design

This is a multi-centre, multi-country, open label, randomised, controlled, adaptive platform trial of antiviral interventions in early symptomatic SARS-CoV-2. The control arm consists of no study drug other than antipyretics. Interventions currently included in the platform are: ivermectin, favipiravir, remdesivir, nitazoxanide, and REGN-CoV-2. We plan to include molnupiravir, nirmatrelvir-ritonavir, Sotrovimab and others depending on local availability and local ethical approval. At each site there is equal allocation to all interventions but with a minimum of 20% of patients randomised to the negative control arm.

#### 2.2 Randomisation

Randomisation is performed via a centralised web-app designed by MORU software engineers using RShiny, hosted on a MORU server. Each study nurse responsible for randomising patients has unique login credentials provided by email from the main study statistician (James Watson). Randomisation sheets are pre-generated for each study site separately using blocks of size K (where K is equal to 3 times the number of interventions available at the site) with an additional 100/K% fuzziness (randomly interchanging one of the allocations per block to avoid the study nurse knowing exactly what the Kth allocation will be).

Randomisation sheets and randomisation event logs are stored on a secure Dropbox folder (professional version that has full version control) which is accessed directly by the RShiny app. Only the study statistician James Watson and the MORU IT manager have read/write privileges to this Dropbox folder. For cross checking purposes, the randomisation app also records the patient age and sex. Each randomisation event is logged with a time-stamp and the identity of the nurse who performed the randomisation. Each time the set of interventions available at a given study site changes, a new randomisation sheet is generated, overwriting the previous one.

#### 2.3 Sample size

The sample size is adaptive (there is no fixed sample size). The required sample size depends on how stringent the thresholds are that define futility or success. We used a simulation approach to determine futility and success thresholds that result in control of the type 1 error at approximately 10% and control of the type 2 error at approximately 20% (see study protocol for exact details). The simulations assumed that the decline in viral loads was log-linear (linear on the cycle threshold (CT) scale), using measurement error estimated from an open access database of prospectively followed individuals with frequent viral load measurement [7, 4]. We calibrated plausible effect sizes using preliminary data from the Regeneron phase 2 studies (this suggested increases of approximately 20% in the slope of the viral clearance on the CT scale) [8]. For these futility and success thresholds, the simulations suggested that the number of patients randomised per arm in order to reach a decision is:

- Approximately 50 patients (average; median is 30) for each efficacious intervention (assuming an effect size of 10% increase in viral clearance slope);

- Approximately 40 patients (average; median is 25) for each inactive intervention (assuming an effect size of 0%).

Therefore, supposing there are 5 interventions of type A in the platform, and assuming that only 1 of the 5 are, in fact, effective, we would need on average a total of 50 + 4*40 = 210 patients randomised to interventions of type A (approximately 66% of the total sample size: a minimum of 20% of samples are randomised to the negative control and thus 13% of patients are randomised to each intervention arm and the positive control arm). This implies a total of 320 patients for the first set of interventions identified (this does not include subsequent patients randomised to the intensive PK sub-studies).

##### 2.4 Framework

All comparisons will be made with respect to the negative control arm (no "across intervention" arm comparisons in the primary analysis). In the case of a site not randomising patients to the negative control (because of local objection), that site will only provide indirect data to support the treatment estimates (under the hierarchical model structure). All decisions concerning efficacy will be based on super-superiority relative to the negative control arm whereby the probability of an effect is defined under the model as the posterior probability that the increase in viral clearance relative to the control arm is greater than 5%.

#### 3 Statistical interim analyses and stopping rules

We plan frequent interim analyses (Figure 1). This is to allow for near-real time monitoring of accrued data in order to detect possible issues in patient recruitment, viral load swabbing, or with the PCR assays at the different sites. We allow for a slightly inflated type 1 error (10%) as this is a phase 2 study (and so the type 1 vs type 2 error trade-off is different from typical phase 3 studies). In addition, meeting the success threshold will trigger a subsequent intensive PK-PD sub-study to determine dose-response effects thus gathering additional efficacy data for that intervention. If there is strong a-priori evidence of antiviral efficacy (e.g. molnupiravir, nirmatrelvir) blood samples for population PK-PD modelling will be obtained from all enrolled patients.

For the type A interventions, we are interested in making decisions regarding futility (highly unlikely that the viral clearance is increased by $> 5\%$) and regarding success (highly likely that the viral clearance is increased by $> 5\%$). For type B interventions (monoclonal antibodies) we are interested in estimating their antiviral effect (in order to interpret and calibrate the observed effect sizes for the type A/C interventions) and detect any temporal changes related to virus genotype (variant escape).

##### 3.1 Interim analysis decision making

Only the study statistician (James Watson) will be unblinded to all interim analyses. Each interim analysis report will sent to the Data Safety and Monitoring Board (DSMB) along with a summary of whether the futility or success thresholds for any of the interventions have been met. The DSMB will then make recommendations for continuing or stopping the intervention arms. Concurrent data from other trials (e.g. safety data) may be used in making these decisions. If an arm is stopped for futility or an intensive PK-PD study is triggered, then all the data pertaining to the negative control arm and the intervention arm will be presented to the Trial Steering Committee (TSC), along with the DSMB recommendations. The interim analysis of the first 50 patients with available qPCR data will be presented unblinded to the TSC in order to determine any necessary final changes to the statistical analysis plan.

##### 3.2 Timing of final analysis

As this is a platform trial, no overall final analysis is planned. For each intervention studied, we define the "final analysis" as:

- For an intervention of type A or C that meets the futility criteria ($<0.1$ probability that viral clearance is increased by at least 5%): final analysis will be timed from when all patients randomised to that arm have passed their day 28 follow-up.

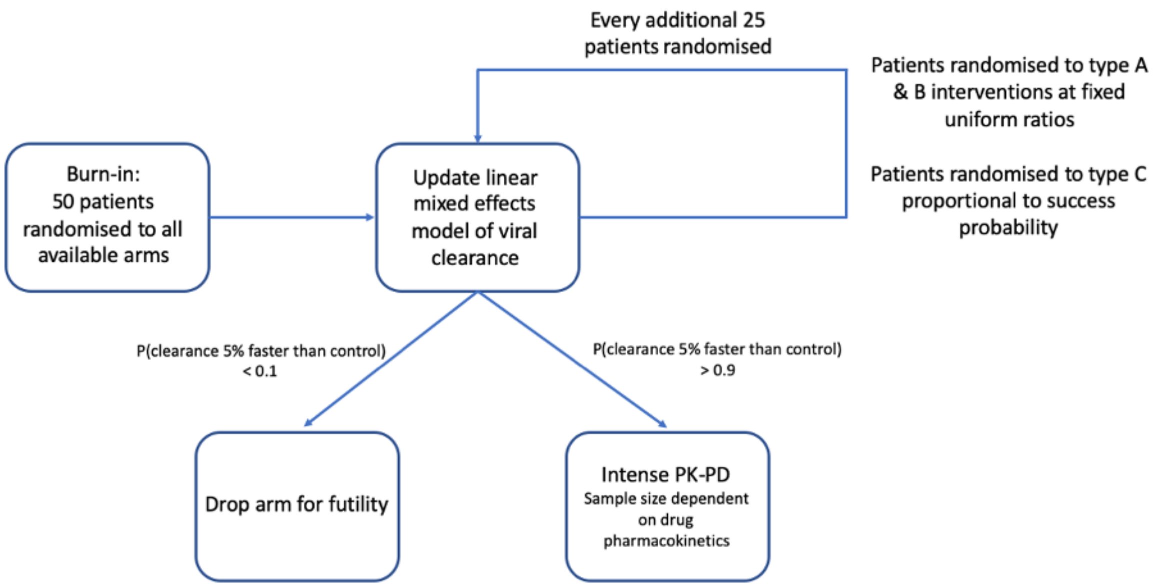

Figure 1: Planned interim analyses and decision rules for stopping an arm or triggering an intensive PK-PD sub-study.

- For an intervention of type A or C that meets the success criteria (>0.9 probability that viral clearance is increased by at least 5%): final analysis will be timed from when the last patient is enrolled in the intensive PK-PD sub-study and has passed their day 28 follow-up.
- For an intervention of type B (monoclonal antibodies), if a decision to drop the arm is made based on loss of efficacy due to variant escape,final analysis will be timed from when the last patient randomized to that arm passes their day 28 follow-up.

##### 3.3 Timing of outcome assessments

For the primary and secondary endpoints we will use all viral load measurements taken up until day 7. Time will be defined as time since randomisation (units of days, including all timepoints $< 8$ days since randomisation as there will be some variation in exact clock times of the follow-up swabs) The tertiary endpoint (all cause hospitalisation) will be taken up until day 28.

#### 4 Statistical Principles

##### 4.1 Posterior estimates

Treatment effects will be estimated under a Bayesian framework using a hierarchical model with weakly informative prior distributions. For the hyperparameters, the prior distributions are chosen for computational reasons to aid model convergence. For the key population level parameters, we will use priors based on analysis of open access data [4]. The prior on the treatment effect is driven by plausible maximum effect sizes given published data on viral decreases in patients randomised to Regeneron or placebo [1].

##### 4.2 Adherence and protocol deviations

This study is open label and thus it is possible that some patients who are randomized to the no study drug arm, or to study interventions perceived to be ineffective, may be given alternative rescue treatment (e.g. the monoclonal antibody Regeneron) if their symptoms persist and the treating physician is worried that they will deteriorate clinically. This can introduce confounding between the treatment allocation and outcome. Notably, if multiple patients randomized to the

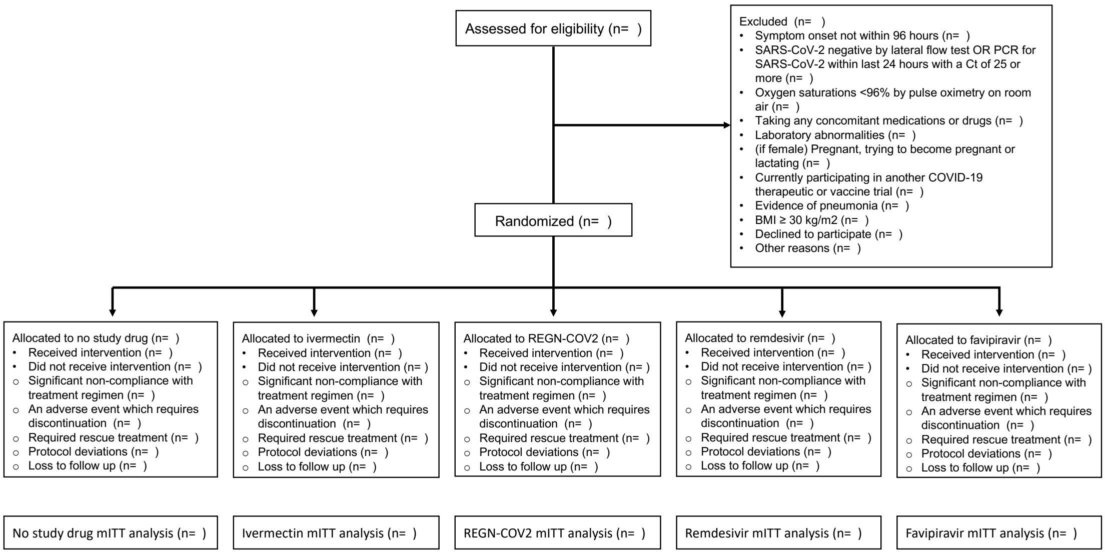

Figure 2: CONSORT diagram summarizing screening, reasons for excluding patients and randomisation.

negative control were in fact given the positive control (e.g. Regeneron) before day 7, this could impact the viral loads during follow-up and thus bias the estimates of the rate of viral clearance in the negative control arm. For all protocol deviations regarding treatment (either stopping treatment for safety reasons or switching treatment arms), we will consider all viral load measurements taken after the deviation as censored, i.e. we will estimate viral load clearance rates only using the data during the period of protocol compliance. We will summarize the number of treatment protocol deviations by site.

#### 4.3 Analysis populations

The primary analysis will be in the intention-to-treat (ITT) population, including follow-up data only from the period of treatment adherence (modified intention-to-treat, mITT). A minimum of 3 days of follow-up data are necessary in order to be included in the mITT population (e.g. patients who discontinue before day 2 will not be included). A secondary analysis will be performed in the per-protocol population, defined as patients who had 100% adherence to the allocated intervention arm. Specific reasons for treatment discontinuation will be outlined and recorded in order to assess confounding bias. The safety population will include all patients who have received at least one dose of the intervention.

### 5 Trial population

Figure 2 shows the CONSORT diagram summarizing the number of patients screened; the reasons for exclusion; the number of enrolled patients randomized to each arm (showing the arms currently open in the Thailand sites); and the number of patients who complete treatment and follow-up and who are included in the modified intention to treat analysis (mITT).

#### 5.1 Screening data

We will summarise the reasons for exclusion for the screened patients not eligible for enrollment. This will not be done for the interim analysis, only for the final analysis once an arm is stopped for futility or success.

##### 5.2 Eligibility

The key trial eligibility criteria are as follows:

- Previously healthy adults, aged 18 to 50 years (low risk of developing severe COVID-19);
- SARS-CoV-2 positive by lateral flow antigen test OR a positive PCR test for SARS-CoV-2 within the last 24 hours with a CT value of less than 25 (all viral targets);
- Symptoms of COVID-19 (including fever, or history of fever) for less than 4 days (96 hours);
- Oxygen saturation $\geq$96% measured by pulse-oximetry at time of screening.

##### 5.3 Withdrawal and follow-up

Level of withdrawal will be tabulated and cover the following aspects:

- Discontinuation from any of the study interventions
- Withdrawal from study follow-up
- Withdrawal from entire study and requests that data is not used.

Data will be tabulated for the timing of withdrawal from follow up or lost to follow up. The number of withdrawals and reason will be recorded. A participant may withdraw from the intervention, or be withdrawn for the following reason.

- Withdrawal of consent by participant
- Alteration of participants circumstances or condition which gives justification for discontinuation as decided by investigator.

A table will summarise the loss to follow up and withdrawals during the study with the corresponding reasons.

##### 5.4 Baseline patient characteristics

The following key baseline characteristics will be summarized, stratified by site:

- Age and sex;
- Baseline viral load (in copies per mL: the mean value of the 4 swabs taken at randomisation);
- Result of SARS-CoV-2 rapid antibody test;
- Baseline quantitative serum antibody titres (when available);
- Vaccination status (vaccine type, number of doses: expressed as not vaccinated, partially vaccinated and fully vaccinated);
- Days since onset of symptoms.

These will be summarised by their mean or median and standard deviation/range for the continuous variables and by their mean for the binary variables.

#### 6 Analysis

##### 6.1 Outcome definitions

The primary outcome is the inferred rate of viral clearance expressed as a slope coefficient (the mean posterior estimate and the 95% credible interval) for the daily change in the log viral load (on the $\Delta$CT scale). This will be inferred under a Bayesian mixed effects (hierarchical) model using the log viral load measurements up until day 7. The primary analysis will be conducted on the observed $\Delta$CT values (40-CT) with batch adjustment.

##### 6.1.1 Viral load quantification

All viral load quantification will be done using ThermoFisher TaqCheck TM SARS-CoV-2 Fast PCR assay. Each PCR assay is run on a 96 well plate, composed of all the aliquots from the transport media of the swabs from 4 patients (10 timepoints each done in duplicate, i.e. 20 samples per patient) and one control set composed of 6 spiked control samples of known viral densities over the range $10^7$ to $10^2$ viral copies per mL, each done in duplicate (12 in total). For each plate (i.e. batch) we can thus estimate a standard curve (which defines the conversion from the CT value to a number of viral copies per mL). At each interim analysis we will perform a quality control check by comparing the estimated standard curves from each plate, both within sites and across sites (each country will run the PCR assays separately). In addition, we will use the standard curve data to estimate heteroscedasticity in the viral load estimation. We expect the PCR measurement error to increase as the viral load decreases. Heteroscedasticity can be shown visually by plotting the true viral load densities in the control duplicated samples against their observed difference in CT value. We can then estimate the variance in the differences as a function of the true viral density.

To help adjust for variation in the human cell content of the swabbed sample the qPCR method assesses viral densities and also human cell densities using RNaseP as a human cell marker.

#### 6.2 Analysis Methods

##### 6.2.1 Statistical model for the analysis of the serial viral load data

The primary analysis will consist of fitting Bayesian hierarchical (mixed effects) linear models to the log viral load data up until day 7. The treatment effect is defined as the proportional change (expressed as a multiplicative term) in the population slope of the daily change in log viral load. We will model the data directly on the $\Delta$CT (40-CT) scale adjusting for batch differences (i.e. each PCR batch has a slightly different standard curve, i.e. transformation to the copies per mL scale). The model jointly analyses the control samples placed on each PCR plate (this allows for inference on the intrinsic measurement error and the batch differences) and the patient samples. The general model likelihood takes the following form:

$$y_{i,t,k,b} \sim \text{Student}\left(\lambda, \alpha_0 + \alpha_i + \alpha_k + \alpha_b + \alpha_{cov} + \gamma x_{i,t} + \beta_0 e^{\beta_k + \beta_i + \beta_{cov} + \beta_{T(i)}} t, \sigma^2_{\text{PCR}} + \sigma^2\right) \quad (1)$$

$$z_{b,d} \sim \text{Normal}\left(\theta_1 + \alpha_b + \theta_2 d, \sigma^2_{\text{PCR}}\right) \quad (2)$$

where:

- $y_{i,t,k,b}$ is the log viral load ($\Delta$CT) for patient $i$ at time $t$ (in days since randomisation) from site $k$, measured from batch $b$.
- $T(i)$ is the randomized treatment allocation for individual $i$.
- $z_{b,d}$ is the $\Delta$CT value for the control sample with known density $d$ on batch (i.e. plate) $b$.
- $\sigma^2_{\text{PCR}}$ is the intrinsic variance in measurement (PCR assay variance).
- $\sigma^2$ is the biological measurement variance.
- $\lambda$ is the degrees of freedom of the student-t error model.
- The slope of viral clearance decomposes into 5 multiplicative terms: the population mean slope $\beta_0$; the site random effect $\beta_k$ ($e^{\beta_k}$ is thus the proportional change in slope); the individual random effect $\beta_i$; the covariate effect $\beta_{cov}$; and the treatment effect (fixed across sites and individuals) $\beta_{T(i)}$.
- The intercept term (baseline log viral load) decomposes into 5 terms: the population intercept $\alpha_0$; the site random effect $\alpha_k$; the individual random effect $\alpha_i$; the batch random effect $\alpha_b$ (PCR assay batch); and the covariate effect $\alpha_{cov}$. All comparisons are made relative to the no antiviral treatment control arm, so we set $\beta_{control} = 0$.
- $x_{i,t}$ is the relative human RNaseP quantification (RNaseP $\Delta$CT value: this is proportional to the log number of human cells) for patient $i$ at time $t$. The parameter $\gamma$ thus provides an adjustment for human cell content in each swab. We scale $x_{i,t}$ so that it has mean 0.

- The parameters $\theta_1, \theta_2$ define the standard curve for the PCR assay, with $\alpha_b$ the same batch effect as for the patient samples (constant shift).

We will fit the data by sequentially fitting five nested models. The base model (model 0: 'vanilla') only includes individual and site random effect terms for the slope and intercept. Model 1 adds the parameter $\gamma$ for the adjustment for human RNaseP. Model 2 adds batch effects, and model 3 adds the control data (control samples with known viral load) allowing for the decomposition of the total variance into the intrinsic machine variance and the biological variance in the patient samples. The final model then incorporates all the covariate adjustment terms $\alpha_{cov}$ and $\beta_{cov}$ (see below for their definition). If analysis only has data from one site, then the site random effect term $\alpha_k$ will be dropped.

We have chosen a student-$t$ distribution for the model likelihood (with the number of degrees of freedom $\lambda$ estimated from the data) as this is robust against departures from normal (Gaussian) error, and against model mis-specification, notably concerning the assumption of log-linear decline in viral loads. Prior analyses carried out in order to prepare this statistical analysis plan suggest that a considerable number (e.g. up to 5%) of viral load densities can depart substantially from the expected distribution under a simple log-linear model with Gaussian error [4]. This can partially be explained by patterns of bi-exponential decay in viral loads. It is unclear if the slope of the second phase of elimination in a bi-exponential decline is affected by interventions. We therefore choose not to fit bi-exponential models as this makes the interpretation of the treatment effect more difficult and prior modelling suggested no major impact in terms of treatment effect estimation [4].

Goodness of fits will be assessed using leave-one-out validation [9] and Bayesian $R^2$ approximation [10].

##### 6.2.2 Covariate adjustment

In addition to the human RNaseP adjustment, the primary analyses will adjust for the following covariates (through the parameters $\alpha_{cov}, \beta_{cov}$):

- The serological status on admission as a binary variable corresponding to the result of the screening rapid antibody test (+/-);
- When available (likely July 2022), quantitative serum antibody at baseline (assay not yet determined);
- Age (in years - scaled to have mean zero);
- A quantitative measure of serological status using a pre-specified antibody (not yet decided). These data will only be available after August 2022.
- Previous SARS-CoV-2 vaccination (as an ordinal variable: 0: no doses; 1: partially vaccinated with only one dose: 2: fully vaccinated with 2 doses; 3: 3 or more doses);
- The SARS-CoV-2 variant of concern (following WHO labels, with the reference strain chosen to be the major Delta lineage which was the dominant variant of concern when the trial started in Bangkok in September 2021; for sub-lineages, e.g. BA.2, data analysis will be exploratory and not part of the primary analysis of treatment effects).
- The trial "epoch": this is defined as the discrete intervals of time at which new interventions enter or exit the study. This is to adjust for temporal effects which could confound across arm comparisons made for non-contemporaneously recruiting arms.

If any of the binary covariates show no variation (e.g. all patients are vaccinated) then we will drop the term from the model. We will fit both the covariate adjusted model and the non-adjusted model in all instances as to check for sensitivity to the covariate model. A sensitivity analysis regarding temporal drift will be done by analyzing each arm individually along with contemporaneously enrolled controls.

#### 6.3 Subgroup analyses

The primary pre-specified subgroup analysis will look at the relationship between the efficacy of the monoclonal antibodies (currently the Regeneron antibody cocktail) and the virus genotype expressed as the major WHO lineages. This will be done by fitting an interaction term $\delta x_{var}(i)e^{\beta_{T(i)}}$ for the relevant $T(i)$; where $x_{var}(i)$ is the virus variant infecting individual $i$.

Secondary subgroup analyses will be carried out only for treatment arms that show an antiviral effect (meet the success criteria). We will look at subgroup effects for (listed in order of priority):

- SARS-CoV-2 variant of concern;
- Serological status on admission (using quantitative serum antibody);
- Vaccination status;
- Age and sex;

#### 6.4 Prior distributions

In all the following, for the normal distributions, the second term corresponds to the scale (standard deviation not the variance).

**Population level parameters**

$$\alpha_0 \sim \text{Normal}(18, 5) \quad \text{(population intercept)}$$

$$\beta_0 \sim \text{Normal}(-2, 2) \quad \text{(population slope)}$$
$$\sigma \sim \text{Normal}(3, 3) \quad \text{(standard deviation of biological noise)}$$
$$\sigma_{\text{PCR}} \sim \text{Normal}(0.5, 1) \quad \text{(standard deviation in assay)}$$
$$\gamma \sim \text{Normal}(0, 1) \quad \text{(human RNaseP adjustment)}$$
$$\beta_{T(i)} \sim \text{Normal}(0, 0.5) \quad \text{log treatment effect}$$
$$\theta_1 \sim \text{Normal}(-3, 3) \quad \text{(standard curve intercept)}$$
$$\theta_2 \sim \text{Normal}(\log_2(10), 1) \quad \text{(standard curve slope)}$$
$$\delta \sim \text{Normal}(0, 1) \quad \text{(variant subgroup effect)}$$

**Covariate effects**

$$\beta_{cov} \sim \text{Normal}(0, 1) \quad \text{(covariate effect on slope)}$$
$$\alpha_{cov} \sim \text{Normal}(0, 1) \quad \text{(covariate effect on intercept)}$$

**Random effects and hyperparameters**

$$\Omega \sim \text{Cholesky}(2) \quad \text{(correlation matrix for individual random effects)}$$

$$\Sigma \sim \text{Exponential}(1) \quad \text{(standard deviation for individual random effects)}$$
$$\alpha_b \sim \text{Normal}(0, \sigma_b) \quad \text{(batch random effects)}$$
$$\sigma_b \sim \text{Exponential}(1) \quad \text{(standard deviation of batch effects)}$$
$$\lambda \sim \text{Exponential}(1) \quad \text{(t-distribution degrees of freedom)}$$

##### 6.5 Sensitivity analyses

Sensitivity to the prior specification will be assessed by multiplying all prior standard deviation terms by 5. Sensitivity to the assumption of log-linear decline will be assessed by fitting an 'up-down' model of viral dynamics [11] (where "up" refers to viral multiplication and "down" the subsequent clearance). Under this model the likelihood is (dropping batch, site, and covariate terms for notational simplicity, these are added analogous to the previous log-linear model) given by:

$$y_{i,t} \sim \text{Student}\left(\lambda, \alpha_0 + \alpha_i + \log\left\{\frac{\beta_1^i + \beta_2^i}{\beta_2^i e^{-\beta_1^i[t-t^i_{\text{max}}]} + \beta_1^i e^{\beta_2^i[t-t^i_{\text{max}}]}}\right\}, \sigma^2_{\text{PCR}} + \sigma^2\right)$$

Under this model, the term $\alpha_0+\alpha_i$ is the individual peak log viral load (on the $\Delta$CT scale) which occurs at time $t^i_{\text{max}}$ (this is the time relative to randomisation, if negative it is therefore unobserved). The parameters $\beta_1^1, \beta_2^i$ are the growth and clearance rates for individual $i$, respectively.

If some patients are enrolled very early in the course of their illness, it is possible that their viral loads increase over the first days of follow-up. This would mean that the clearance rate estimated under a simple log-linear decline model would be biased towards a shallower slope. Randomisation protects against confounding bias as the sample size increases, however for small sample sizes unequal allocation of 'pre-peak' versus 'post-peak' individuals could influence treatment effect estimates. Treatment effects are inferred in the same way as for the log-linear model (proportional change in the population slope coefficients), by acting on $\beta_2$ (note that in reality, the treatment effect most likely would change all three parameters $\alpha_0, \beta_1, \beta_2$, however this would mean inferring three separate treatment effects as the model is non-mechanistic).

##### 6.6 Missing data

Missing viral load will not be imputed. The model will be fit to all available viral load densities up until day 7 or day of treatment deviation. For all analyses, missing data on the key baseline variables (vaccination, virus genotype, antibody rapid test) will be imputed at random using the observed site-specific covariate distribution multiple times (5 times, the model will be refit). We do not expect much missing data (eg $<1\%$). If delays in genotyping result in missing variant data, we will impute the variant using genetic surveillance prevalence data.

##### 6.7 Harms

Safety analyses will include all patients who have received at least one dose of the intervention. The safety and tolerability data will be pooled from all the sites that receive the same study intervention. The safety and tolerability will be assessed by comparing the frequency of adverse events and serious adverse events when compared to the control intervention.

All adverse event summaries will refer to treatment emergent adverse events, i.e. adverse events that newly started or increased in intensity after study drug administration (or from hour 0 in the control group receiving no intervention). Adverse events will be graded according to CTCAE V5.0. Adverse Events (AEs) of grade 3 and above will be recorded, the grading is as follows - 1 = mild, 2 = moderate, 3 = severe, 4 = life-threatening, 5 = fatal. All Serious Adverse Events (SAEs) will be recorded, a serious adverse event was any untoward medical occurrence that resulted in death, was life threatening, required inpatient hospitalization or prolongation of existing hospitalization, resulted in persistent or significant disability/incapacity or consisted of a congenital anomaly or birth defect.

We will tabulate AE of grade 3 and higher, SAEs, and deaths, by treatment arm with comparisons made relative to the no study drug arm.

None of the drugs evaluated will be pre-registration or pre-approval, so adverse effect profiles for each are already well known.

##### 6.8 Statistical software and analysis implementation

All analyses will be be in done in R. Interim analyses and the final analyses will be done using RMarkdown scripts, pre-coded to ensure full transparency. All code for the statistical analysis is available on the following github repository: `https://github.com/jwatowatson/PLATCOV-SAP`. This ensures full version control by tracking all changes made to the analysis code.

The statistical models for the analysis of the serial viral load data are written in *stan* and fitted via the *rstan* interface [12]. These are available on the github repository. For each model, we will run 6 parallel chains for 10,000 iterations, discarding half for burn-in and thinning every 40 (thus 1,000 posterior draws).

Convergence of Bayesian fits will be assessed visually by examining the traceplots of the key model parameters and assessing the Rhat values for each parameter (Gelman-Rubin statistic).
